# Quantitative SARS-CoV-2 viral-load curves in paired saliva and nasal swabs inform appropriate respiratory sampling site and analytical test sensitivity required for earliest viral detection

**DOI:** 10.1101/2021.04.02.21254771

**Authors:** Emily S. Savela, Alexander Winnett, Anna E. Romano, Michael K. Porter, Natasha Shelby, Reid Akana, Jenny Ji, Matthew M. Cooper, Noah W. Schlenker, Jessica A. Reyes, Alyssa M. Carter, Jacob T. Barlow, Colten Tognazzini, Matthew Feaster, Ying-Ying Goh, Rustem F. Ismagilov

## Abstract

Early detection of SARS-CoV-2 infection is critical to reduce asymptomatic and pre-symptomatic transmission, curb the spread of variants by travelers, and maximize treatment efficacy. Low-sensitivity nasal-swab testing (antigen and some nucleic-acid-amplification tests) is commonly used for surveillance and symptomatic testing, but the ability of low-sensitivity nasal-swab tests to detect the earliest stages of infection has not been established. In this case-ascertained study, initially-SARS-CoV-2-negative household contacts of individuals diagnosed with COVID-19 prospectively self-collected paired anterior-nares nasal-swab and saliva samples twice daily for viral-load quantification by high-sensitivity RT-qPCR and digital-RT-PCR assays. We captured viral-load profiles from the incidence of infection for seven individuals and compared diagnostic sensitivities between respiratory sites. Among unvaccinated persons, high-sensitivity saliva testing detected infection up to 4.5 days before viral loads in nasal swabs reached the limit of detection of low-sensitivity nasal-swab tests. For most participants, nasal swabs reached higher peak viral loads than saliva, but were undetectable or at lower loads during the first few days of infection. High-sensitivity saliva testing was most reliable for earliest detection. Our study illustrates the value of acquiring early (within hours after a negative high-sensitivity test) viral-load profiles to guide the appropriate analytical sensitivity and respiratory site for detecting earliest infections. Such data are challenging to acquire but critical to design optimal testing strategies in the current pandemic and will be required for responding to future viral pandemics. As new variants and viruses emerge, up-to-date data on viral kinetics are necessary to adjust testing strategies for reliable early detection of infections.

## Introduction

Early detection of SARS-CoV-2 infection is needed to reduce asymptomatic and pre-symptomatic transmission, including the introduction and spread of new viral variants from travelers. More than half of transmission events^1^ occur from pre-symptomatic or asymptomatic persons. Early detection enables individuals to self-isolate sooner, reducing transmission within households and local communities, and to vulnerable populations, including individuals hospitalized for non-COVID-19 illnesses and individuals at high risk for severe disease due to multiple medical comorbidities (e.g., residents of skilled nursing or long-term-care facilities or memory-care facilities). Low-sensitivity nasal-swab tests are commonly used for SARS-CoV-2 detection and symptomatic testing.^2^ As new variants-of-concern emerge, e.g. the Delta variant with increased transmissibility,^3–5^ high viral loads,^4,6^ and outbreaks with large numbers of breakthrough infections,^7^ it is clear that testing strategies (analytical sensitivity and sample type) need to be adjusted to diagnose infections earlier.

Although national vaccination efforts are reducing severe COVID-19 outcomes in the U.S., a sizable portion of the world’s population is likely to remain unvaccinated due to limited vaccine availability, medical ineligibility (in the U.S., children under 12 years of age are not yet eligible), or personal preference. Thus, testing remains an important tool for preventing outbreaks among children in schools and daycare facilities (where children under age 2 cannot wear masks), which may spread to the community and increase rates of infection among high-risk and unvaccinated individuals. Tests that detect early infections are also important to prevent viral transmission in congregate settings with high-risk or unvaccinated populations, such as hospitals, college dormitories, homeless shelters, correctional facilities, summer camps for children, elementary schools, and long-term care facilities.

Beyond outbreak prevention and control, early detection of COVID-19 may also be useful for individual patient care, as high-risk patients who are identified early can be monitored and treatment initiated swiftly if it becomes appropriate. Several treatments show exclusive or increased efficacy only when given early in the infection. The advantage of earlier treatment initiation is likely due to reduction of viral replication either directly or by promotion of an early effective immune response, which prevents a later exaggerated inflammatory response.^8^ Results of the ACTT-1 trial demonstrated a survival benefit in patients for whom Remdesivir was initiated in the early stages of treatment (supplemental oxygen only), but that benefit was lost once disease progressed, and advanced respiratory support was needed.^8–10^ Convalescent plasma failed to show efficacy in a study where the median time to entry in the study was 8 days after symptom onset^11^ but demonstrated protection against progression to respiratory failure when given to individuals of advanced age earlier in the course of the illness.^12^ Similarly, the use of anti-SARS-CoV-2 monoclonal antibody therapy (bamlanivimab or casirivimab plus imdevimab) did not show benefit over placebo in a cohort of hospitalized patients.^13^ However, when given to outpatients with mild or moderate COVID-19, who may have otherwise progressed to hospitalization later in the course of illness, reductions in emergency room or medical visit rates and more rapid declines in viral load^14–16^ have been observed. Further, a greater effect was observed among the subgroup of patients who had not yet developed a detectable endogenous antibody response.^8,15^

However, it is currently unclear which testing strategy can detect SARS-CoV-2 infection at the earliest stages. Does one need a high-sensitivity test, or would a low-sensitivity test suffice? Which sample type should one use?

Tests with high analytical sensitivity can detect low levels of molecular components of the virus (e.g. RNA or proteins), in a sample. Analytical sensitivity is described by the limit of detection (LOD) of a test (defined as the lowest concentration of the viral molecules that produces 95% or better probability of detection). The lower the LOD, the higher the analytical sensitivity of the assay. LOD of SARS-CoV-2 diagnostic tests are described in various units; the most directly comparable among tests are units that report the number of viruses (viral particles) or viral RNA copies per milliliter of sample. Viral RNA copies/mL are roughly equivalent to genome copy equivalents/mL (GCE/mL) or nucleic acid detectable units/mL (NDU/mL). These LOD values are tabulated by the U.S. Food & Drug Administration (FDA).^17^ High-sensitivity tests have LOD values equivalent to ∼10^2^ to 10^3^ copies/mL of sample, whereas low-sensitivity tests have LOD values equivalent to ∼10^5^ to 10^7^ copies/mL. Therefore, to choose the appropriate test for reliable early detection, one needs to measure viral loads present in samples collected early in the course of infection,^18^ and then choose a test with an LOD below that viral load. Initial data by us^19^ and others^20,21^ show that, at least in some humans, SARS-CoV-2 viral load can be low (in the range of 10^3^–10^5^ copies per mL of saliva sample) early in infection, therefore only high-sensitivity tests would reliably detect infection.

Sampling site or specimen type may also be critical to early detection. Other respiratory viruses have been shown to have detection rates that vary by sampling site,^22^ which have occasionally been linked to viral tropism. For example, the cellular receptor for entry of MERS-CoV is expressed nearly exclusively in the lower respiratory tract, prompting recommendations for diagnostic testing of specific sample types (bronchoalveolar lavage, sputum and tracheal aspirates).^23^ A previous study on SARS-CoV found high levels of viral RNA in saliva and throat wash early in the infection course (before the development of lung lesions), to suggest saliva as a promising sample type for early detection.^24^ Although nasopharyngeal (NP) swab is often considered the gold standard for SARS-CoV-2 detection, it requires collection by a healthcare worker and is not well tolerated. Furthermore, the performance of NP swabs for early detection of current SARS-CoV-2 variants is not known. Other sample types, such as nasal (anterior-nares or mid-turbinate) swabs^25–28^ and saliva^29–32^ are more practical, especially for repeated sampling in serial surveillance testing (also described as “screening”).

Studies comparing paired samples (collected from different locations in the respiratory tract) from the same individual are inconsistent in their findings of which sample type had the better sensitivity. Some studies concluded that nasal swabs outperform saliva/oral fluid^27,33–36^ and had higher viral loads,^26,28,34^ whereas others concluded that testing performance in different locations of the respiratory tract is similar.^30,37–43^ Some studies have observed detection of SARS-CoV-2 in saliva before nasal swabs, or in saliva but not in nasal swabs;^21,44,45^ one study of an emerging variant (B.1.616) showed poor detectability of SARS-CoV-2 in NP swabs.^46^ It is not known whether NP is the best respiratory site for currently circulating variants and emerging variants.

There are several possible explanations for these inconsistencies in respiratory sampling site. Most studies comparing clinical sensitivity of different respiratory sites for SARS-CoV-2 detection focused only on viral detection, not viral-load quantification, which is needed to infer whether detection would have been achieved by assays with different LODs. Although a few studies collected samples in RNA-stabilizing buffers,^40,47–49^ most have collected dry-swabs or saliva in sterile collection vessels;^26–28,30,37,50–57^ without an RNA-stabilizing buffer, introducing risk of viral degradation during transport and handling, which will affect detection and quantification. Most studies that compared multiple respiratory sites for SARS-CoV-2 detection selected individuals already known to be positive for SARS-CoV-2, thus missing the very earliest detectable loads and also not having context for how far along the course of infection that individual might have been. An excellent study comparing nasal swabs and saliva sampling early in the infection among adults at a university^58^ suggested that high-sensitivity testing is needed for early detection; by leveraging university saliva surveillance testing and enrolling close contacts, the researchers reported longitudinal viral kinetics data from 60 participants, 3 of whom (based on our interpretation of the study) were negative in both sample types upon enrollment, allowing for definitive quantification of the earliest day(s) of infection. The thorough work done here shows both how important it is to obtain early samples, but also how difficult it is to capture samples from which to assess the earliest days of infection.

Negative samples preceding the first positive result are needed confirm with high resolution the true starting point of longitudinal measurements on viral load. In order to compare diagnostic performance at different stages of the infection, studies with longitudinal data often align the comparisons to an infection time point—typically days relative to symptom onset,^27,30,35,47,58–64^ laboratory diagnosis (i.e., first positive test result),^27,38,59,65^ or peak viral load,^58^ but these measures can be highly variable among people. Misalignment of illness stage may confound comparisons of sample type sensitivity or viral loads throughout the course of infection. Despite the urgency of defining optimal diagnostic strategies to contain further outbreaks (and spread of variants-of-concern), there is a lack of quantitative data on longitudinal SARS-CoV-2 viral load in paired sample types with sample collection starting prior to earliest detectable viral loads.

To understand the required test sensitivity and the optimal sample type for earliest SARS-CoV-2 detection, we designed a case-ascertained study of household transmission with high-frequency sampling of both saliva and anterior-nares nasal swabs. Building on our earlier work,^19^ we enrolled individuals from Los Angeles County, California, ages 6 and older who had recently tested positive (household index case), and their exposed household contacts at risk of infection. All participants self-collected saliva and anterior-nares nasal swabs twice daily, in the morning upon waking and before bed. Importantly, all samples were immediately placed in a guanidinium-based inactivating solution (see Methods) that preserves viral RNA. We measured the stability of RNA in this buffer over the time periods relevant to our sample processing. Samples were screened for SARS-CoV-2 *N1* and *N2* gene positivity using a high-sensitivity assay and if a transmission event was observed (a previously SARS-CoV-2 negative participant tested positive in at least one sample type), we quantified viral loads in all samples (saliva and nasal swab) prospectively collected from that participant for at least two weeks from their first positive. Quantification was performed via quantitative reverse-transcription PCR (RT-qPCR), with a subset of measurements validated by reverse-transcription droplet digital PCR (RT-ddPCR), capturing the early and full course of acute SARS-CoV-2 infection with high sensitivity.

## Results

First, we established and validated two independent quantitative assays to measure SARS-CoV-2 viral load: a RT-qPCR based on the assay put forth by the U.S. Centers for Disease Control and Prevention (CDC)^66^ and a RT-ddPCR assay developed by Bio-Rad.^67^ Both of these assays received an emergency use authorization (EUA) for qualitative, but not quantitative, detection of SARS-CoV-2. In initial testing, when combined with standard KingFisher MagMax sample preparation protocols, these assays performed well to quantify heat-inactivated SARS-CoV-2 viral particles spiked into commercially available SARS-CoV-2 negative saliva and nasal fluid from pooled donors. However, they did not provide reliable quantification when we analyzed individual saliva samples freshly collected from positive participants in this study. Carryover of materials from some of the mucus-rich samples was inhibitory, as determined by RT-ddPCR analysis of dilutions of eluted RNA (data not shown). We optimized the extraction and each quantitative assay protocol (see Methods) to obtain more reliable quantification of SARS-CoV-2 viral load. We confirmed that the LOD of the modified assay was 1,000 copies/mL or better (see Methods, Fig. S1).

We cross-validated our quantification methods in two steps. First, we used commercial, heat-inactivated SARS-CoV-2 viral particles to establish calibration curves for both saliva and swab samples to convert RT-qPCR quantification cycle values (Cq, also referred to as cycle thresholds, Ct) to viral load. Input particle concentrations for each point on the curve were calculated based on the stock quantification reported on the certificate of analysis for each lot of particles. We could not extend the calibration curve to very high viral loads because of the limited concentration of viral particle stock; so, to confirm performance at high viral loads, we quantified 42 swab and 63 saliva samples from SARS-CoV-2-positive participants with both RT-qPCR and RT-ddPCR methods. We observed excellent concordance between the calibration curve (Data in Fig. S2), RT-qPCR and RT-ddPCR assays over the entire dynamic range of input concentrations (Fig. 1), even though RT-qPCR eluents were run as-is and RT-ddPCR eluents from high-concentration samples were significantly diluted. For nasal-swab samples, RT-ddPCR values were slightly below the RT-qPCR values, however this difference was consistent across the entire dynamic range, indicating no concentration-dependent biases like enzymatic inhibition. We chose not to adjust the calibration curve to fit RT-ddPCR values; we reported the concentrations based on the calibration curves derived from the certificate of analysis from the BEI reference material. For saliva samples, all points tightly clustered around the x=y line.

**Figure 1.**
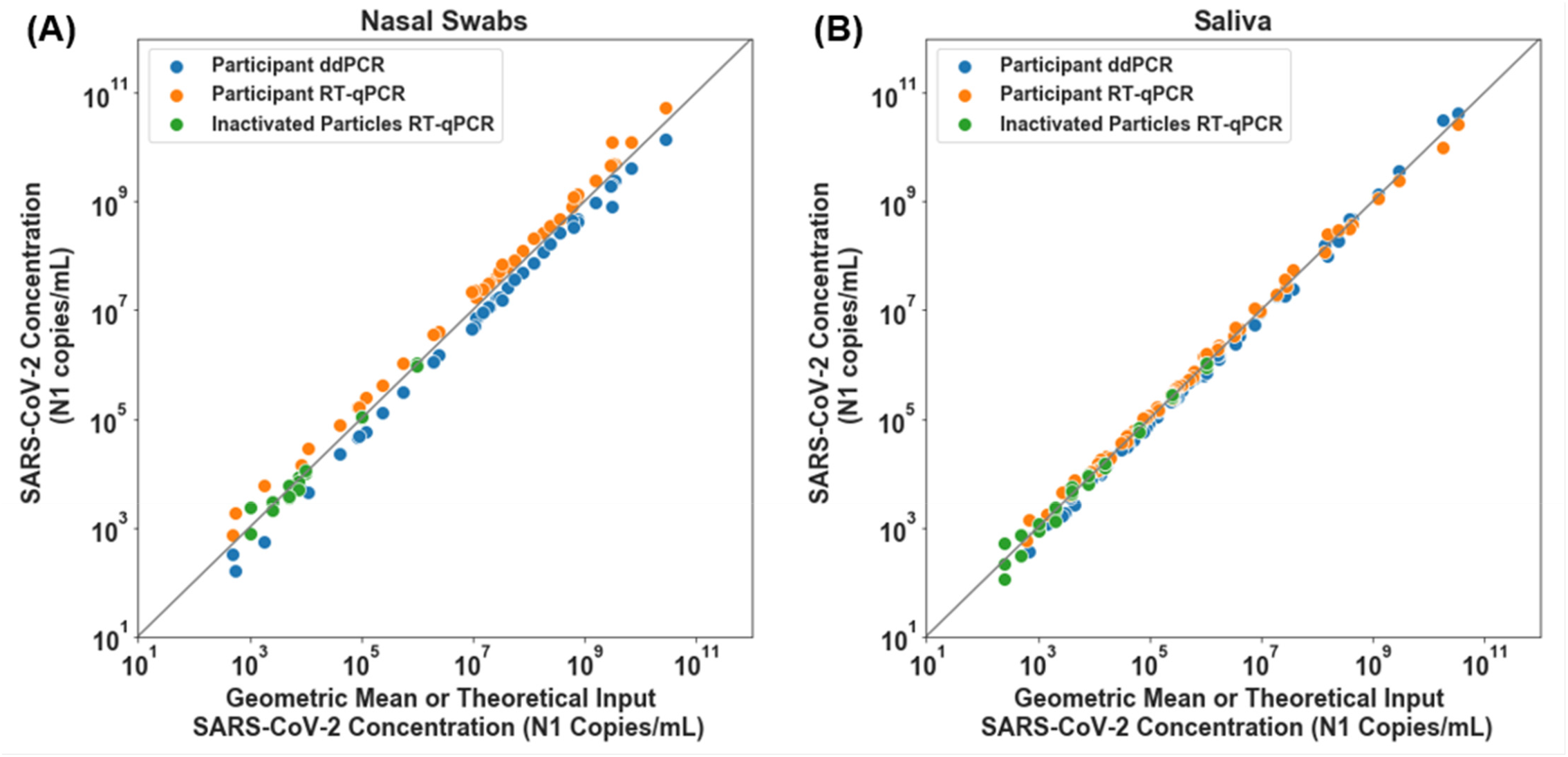
SARS-CoV-2 viral load quantification for nasal-swab (A) and saliva (B) specimens from positive participants measured with RT-ddPCR and RT-qPCR. Participant nasal swab (A) or saliva (B) SARS-CoV-2 *N1* concentration (copies/mL) per detection method, RT-ddPCR (Blue circles) and RT-qPCR (orange circles) plotted against geometric mean of RT-qPCR and RT-ddPCR viral load concentrations. A total of 42 nasal swab and 63 saliva samples from study participants were quantified with both methods. Theoretical SARS-CoV-2 concentration input represents data from calibration curves created with a dilution series of contrived samples prepared using commercial, inactivated SARS-CoV-2 particles spiked into commercially available SARS-CoV-2 negative saliva or nasal fluid pooled from human donors (green circles), extracted and detected with RT-qPCR. Grey line represents x=y.

Next, to quantify viral load at the earliest stage of infection, we analyzed the viral loads in the saliva and nasal swabs of participants who were negative in both sample types upon enrollment and became positive during their participation in the study (Fig. 2). We extended each participant’s enrollment in our study to acquire 14 days of paired saliva and nasal-swab samples starting from the first positive sample. Data in Fig. 2 reports the viral load concentrations as measured on the day of extraction. All samples were stored at 4 °C before extraction; time of storage varied between 0-27 days. The stability of SARS-CoV-2 RNA in nasal-swab samples was slightly lower (1 Cq loss of RNA detected after a median of 15 days) than the stability of SARS-CoV-2 RNA in saliva samples (1 Cq loss of RNA detected after a median of 51 days) (Fig. S3). An assessment of how viral-load measurements in Fig. 2 may have been affected by time between sample collection and quantification is included in Fig. S4. Given the large dynamic range of the viral loads in these samples (∼24 Cq or about 10,000,000 fold), we considered stability corresponding to a 1 Cq (2 fold) loss to be adequate.

**Figure 2.**
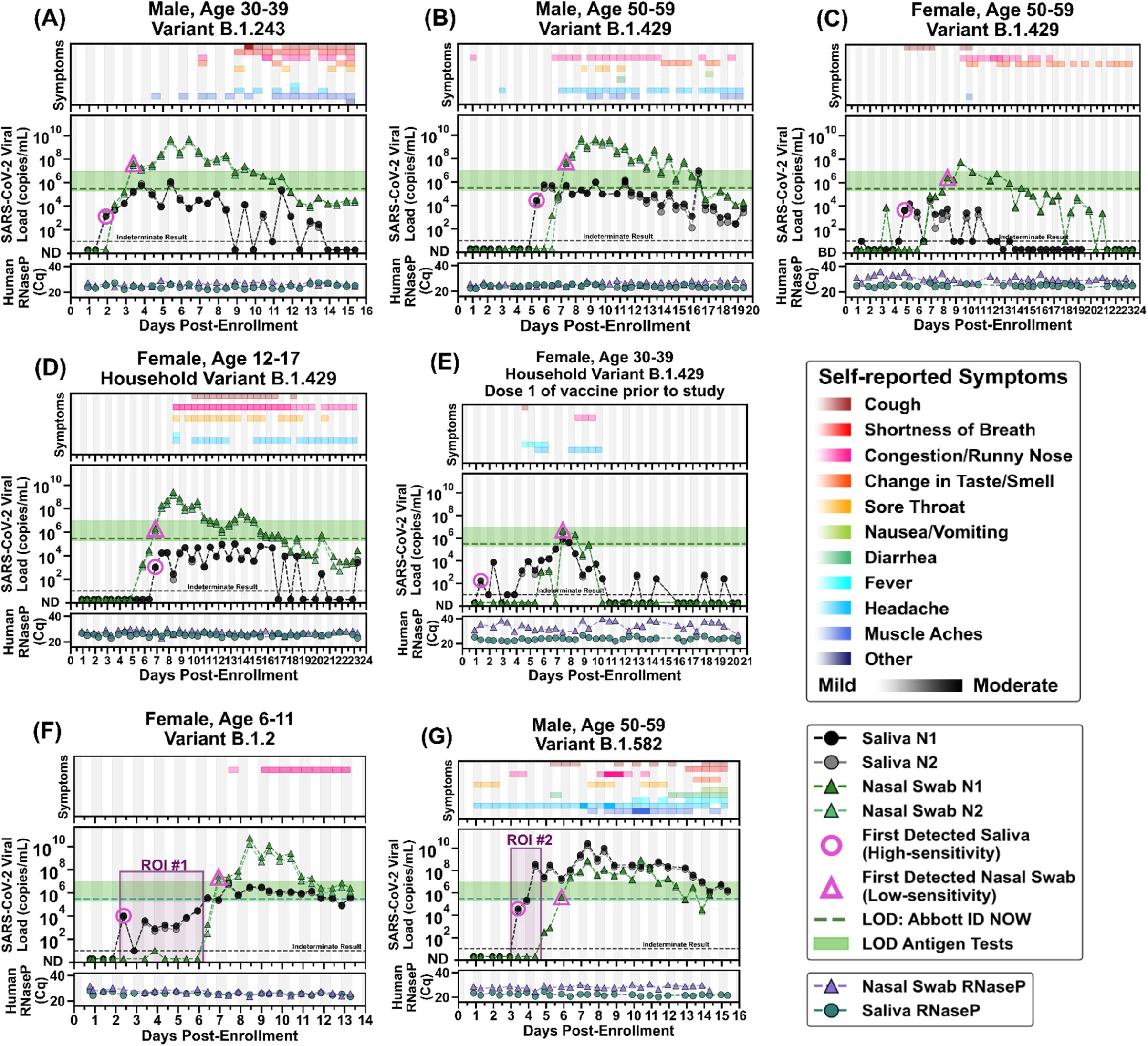
Symptoms and SARS-CoV-2 viral loads in paired saliva and nasal-swab samples of seven participants who became SARS-CoV-2 positive during study participation. **(A-G)** Self-reported twice-daily symptom data over the course of enrollment are shown as a top panel for each of the participants (see color-coded legend for symptom categories). Demographic data including any reported medical conditions are included in Table S1. Viral loads are reported for the *N1* and *N2* genes of SARS-CoV-2 for both saliva (black and grey circles) and nasal-swab samples (dark-green and light-green triangles); ND = not detected for Cqs ≥40; Samples with an indeterminate result by the CDC RT-qPCR assay are shown along the horizontal black dashed line. (see Methods for details). The limit of detection (LOD) of the Abbott ID NOW (300,000 NDU/mL^17^) is indicated by the horizontal green dashed line; the range of LODs of antigen tests (horizontal green bar) are shown for reference (data are from Table S2 in ref. ^19^). A diagnostic test does not provide reliable detection for samples with viral loads below its LOD. For each participant, the first detected saliva point is emphasized with a pink circle and their first nasal-swab point above the LOD of the ID NOW is emphasized with a pink triangle. Vertical shading in grey indicates nighttime (8pm – 8am). Internal control of *RNase P* gene Cqs from the CDC primer set are provided for each sample to compare self-sampling consistency and sample integrity (failed samples, where *RNase P* Cq ≥40, are not plotted). Participant gender, age range, and SARS-CoV-2 variant are given in each panel’s title. Two regions of interest (ROI) are indicated by purple-shaded rectangles and discussed in the main text.

Here we report complete viral load curves in saliva and anterior nares nasal swabs from seven individuals (Fig. 2). Each of these participants tested negative (ND, not detected; Fig. 2) in both saliva and nasal swabs upon study enrollment, ensuring that we capture the earliest days of infection. *RNase P* Cq values remained consistent throughout the collection period for both saliva and nasal swabs for most of the participants (Fig. 2A, B, D, F, and G), indicating observed changes in viral loads were likely not a sampling artifact but rather reflected the underlying biology of the infection. Because nasal swabs are commonly used with low-sensitivity tests, and because such tests are proposed to be utilized for SARS-CoV-2 serial surveillance testing (screening),^68,69^ we wished to compare whether low-sensitivity testing with nasal swabs could provide equivalent performance to high-sensitivity testing with saliva.^31,52,70^

In six out of seven participants, high-sensitivity saliva testing would have been superior for early detection of SARS-CoV-2 infection compared with low-sensitivity nasal-swab measurements (and equivalent for the seventh participant). In the first participant, (Fig. 2A), detection occurred first in saliva at low viral load (1.3×10^3^ copies/mL *N1* gene, pink circle), while the nasal swab remained negative, and days before the participant reported any symptoms. As measured, viral load in nasal-swab samples reached the level of LOD of low-sensitivity tests 1.5 days after the first saliva positive samples (pink triangle). This same pattern of earlier detection in high-sensitivity saliva was observed in five of the other six participants: high-sensitivity saliva was 2.5 days earlier (Fig. 2B), 3.5 days earlier (Fig. 2C), 6 days earlier (Fig. 2E), 4.5 days earlier (Fig. 2F), and 2.5 days earlier (Fig. 2G). Even conservatively accounting for potential decreases of viral RNA in the nasal swab resulting from delays between sample collection and quantification only impact the interpretation of two points, conservatively decreasing the delay from 1.5 to 1 day for the first participant (Fig. 2A and Fig. S4A) and from 3.5 to 3 days for the third participant (Fig. 2C and Fig. S4C). The maximum delay in detection between saliva and nasal swab in an unvaccinated person was observed by the youngest participant in our study (see ROI#1 of Fig. 2F). This participant had detectable but low viral load (10^3^-10^4^) of SARS-CoV-2 RNA in saliva for 4 days while nasal swabs remained negative by high-sensitivity measurements. The nasal viral load spiked above 10^10^ copies/mL. Even after spiking to high viral load in nasal swab, her only symptoms were mild congestion/runny nose. Even with high-sensitivity nasal swab testing, only one participant tested positive in nasal swab before saliva (Fig. 2D). In this participant, SARS-CoV-2 RNA was detectable with a high-sensitivity nasal swab 1 day before high-sensitivity saliva. Nasal swabs reached the detection range of low-sensitivity tests (pink triangle) on the same day as the first saliva sample was detected (pink circle). For all seven participants, high-sensitivity saliva testing would have detected SARS-CoV-2 RNA either the same day or up to 6 days before viral loads in nasal swab reached the detection limits of low-sensitivity nasal swab tests. This pattern of earlier positivity but lower viral loads in saliva compared with nasal swabs was not due to RNA degradation in saliva; when we examined the potential effect of RNA stability there was little effect on RNA concentration over time in saliva samples stored in the preservation buffer at 4 °C (Fig. S3, Fig. S4).

Three participants (Fig. 2C–E) were infected with the same variant, B.1.429 (CAL20), classified as a variant-of-concern at the time of this study. The SARS-CoV-2 variant for the participants in Fig. 2D and Fig. 2E were inferred from the sequenced sample of the household’s presumed index case. Saliva viral loads for each of these participants (Fig. 2C-E) were low. Of note, the participants in Fig. 2C and 2E showed high *RNase P* Cq values (indicating low concentration of the human control target); and variability of *RNase P* Cq values across the nasal-swab samples suggests that inconsistent swab-sampling quality could have impacted these participants’ viral load data, and should be taken into account when interpreting those data. These two participants (Fig. 2C and 2E) also had low viral load in both saliva and nasal swabs. Their viral load measurements were near the LOD of our assay, and therefore as expected, many samples from these participants had indeterminate results.

The fifth participant (Fig. 2E) had received one dose of the Pfizer-BioNTech COVID-19 vaccine^71^ 13 days prior to her first sample. As the only participant in our study who had received a vaccine dose, observations here are not powered to make conclusions about viral load due to vaccination. This participant had very low viral loads in both saliva and nasal-swab sample types and several indeterminate test results when adjacent viral load measurements were near the LOD of our assay. Peak viral loads were lower and shorter in duration compared with the other six study participants. A high-sensitivity saliva test (pink circle) detected the infection 6 days before viral loads reached the lower range of the LODs of low-sensitivity tests (pink triangle) for this participant. Recent data from individuals infected by the Delta variant suggest viral loads in breakthrough infections are not impacted by vaccination status.^7,72^

The final participant (Fig. 2G) with medical history significant for obesity, reported experiencing symptoms beginning 3 days prior to enrollment, and tested negative for SARS-CoV-2 by a CLIA-lab test 2 days prior to enrollment in the study. This person would later report more diverse symptoms (including gastrointestinal symptoms) and with higher symptom-severity ratings than the other six participants. Remarkably (see ROI#2 in Fig. 2G), saliva viral load spiked to 3.7×10^8^ viral copies/mL (*N1* gene target) while SARS-CoV-2 RNA remained undetectable in nasal swab, even by the high-sensitivity assay used here. This contrast between high and likely infectious viral load in saliva^73^ at the same time point as a negative nasal swab emphasizes the need for careful choice of sampling site and test sensitivity in the early stages of SARS-CoV-2 infection to minimize transmission.

Compiled data from all seven participants highlights the non-trivial interplay of anatomical sampling site, infection stage (early vs late), and diagnostic test sensitivity (Fig. 3). Participant results were aligned to the first positive result from either sample type (day 0) and the percentage of positive tests was calculated for each time point (0.5-day intervals) from the first positive sample (Fig. 3A). Based on SARS-CoV-2 RNA viral loads, we found that high-sensitivity saliva testing (100% of participants detected) outperforms low-sensitivity nasal swabs (86% of participants detected) at 5.5 days of positivity (Fig. 3A). If we compare high-sensitivity saliva testing to high-sensitivity nasal-swab testing, this difference is 4.0 days (Fig. S5). However, we emphasize that our data paint a more nuanced view than “saliva is better than swab.” Analytical sensitivity of the test strongly impacts the overall test performance and the preferred sample type: Low-sensitivity saliva testing would have likely yielded the most false-negatives in the first days of infection (only 3 of 7 participants after 5.5 days of positivity) and detecting at most 60% of participants (Fig. S5).

**Figure 3.**
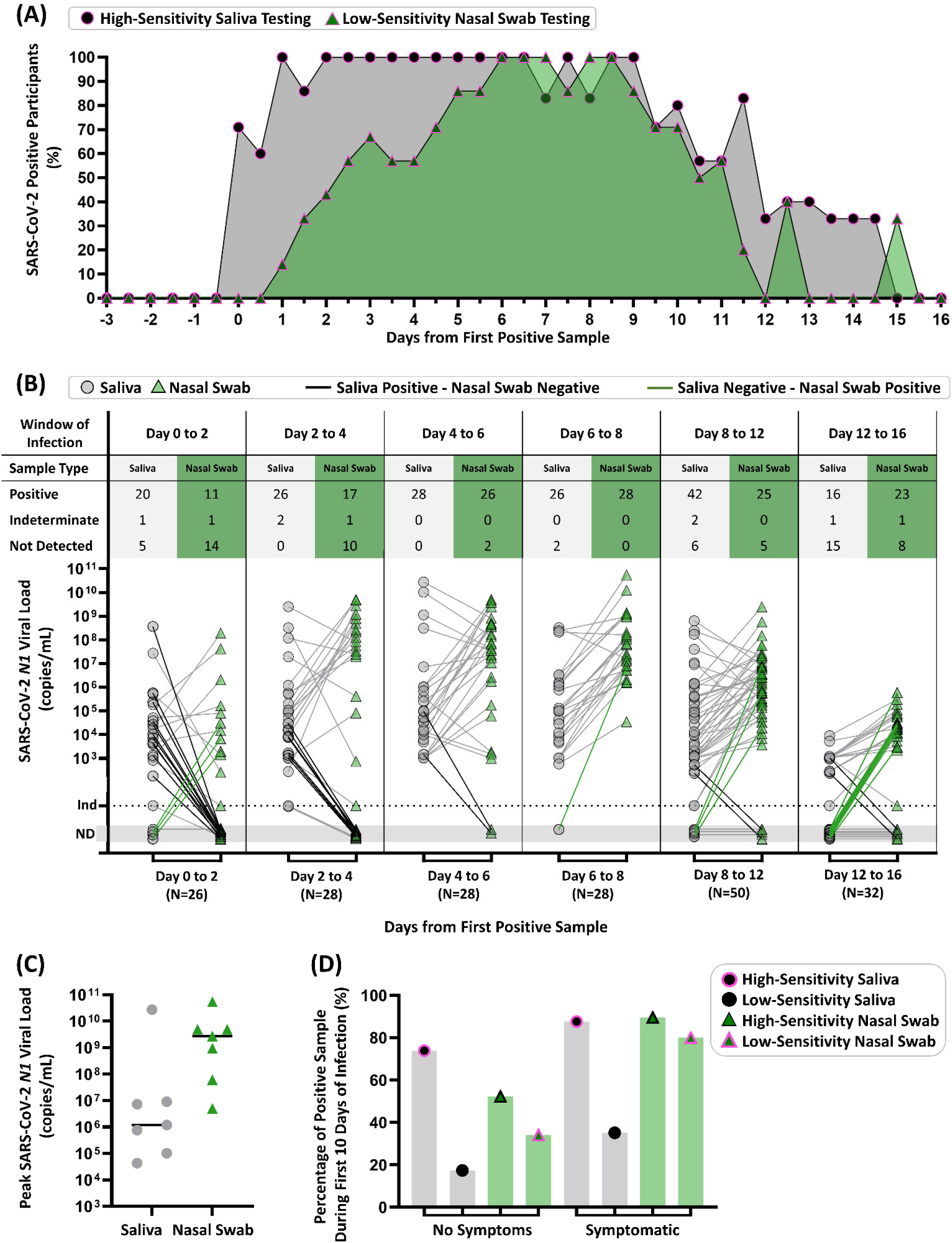
Summary of Diagnostic Insights from Study Participants who became infected with SARS-CoV-2 while enrolled in the study. **(A)** Participant infection time courses were aligned to the first high-sensitivity (LOD of ≤1 × 10^3^ copies/mL) positive result from either saliva or nasal swab sample type (day 0) and the percentage of positive tests was calculated for each time point (0.5-day intervals) from the first positive sample. The predicted performance of low-sensitivity nasal swabs was determined using the individual SARS-CoV-2 *N1* viral load values for each participant individually shown in Fig. 2, or above a viral load threshold of 1.9×10^5^ copies/mL, which is the most-sensitive of the low-sensitivity tests we included in shaded region of Fig. 2. We show the percentage of participants who were detected by our high sensitivity saliva test (black circle) or could be inferred to be detectable by a low-sensitivity nasal swab test (green triangle) at a given timepoint. **(B)** Quantitative SARS-CoV-2 *N1* viral loads of paired samples collected during time windows of the infection (aligned to first positive result by high sensitivity testing of either sample type) are shown for saliva (grey circles) and nasal swabs (green triangles). Paired samples for a given time point are connected with grey lines, with emphasis on paired samples where only saliva (black connecting line) or nasal swab (green connecting line) were positive. ND = Not Detected, Ind = Indeterminate result. **(C)** Peak SARS-CoV-2 *N1* viral loads measured in saliva (grey circles) and nasal swab (green triangles) for each of the seven participants are shown. Horizontal black line indicates the median. **(D)** Percentage of positive test results (out of total number of tests) are shown for the first 10 days for each participant (day 0 corresponds to the first positive test result in either sample type). Saliva (grey bars) and nasal swab (green bars) are shown. Positivity was either observed (by our high sensitivity test) or inferred to be positive by viral loads above a LOD threshold of 1.9e5 copies/mL for low-sensitivity tests. The symptomatic category includes any sample where one or more symptom was reported at the time of sample collection.

Next, we plotted viral loads in each respiratory site starting from the first positive test (Fig. 3B). Paired samples for a given time point are connected with grey lines, with emphasis on paired samples where only saliva (black connecting line) or nasal swab (green connecting line) were positive. From day 0 to day 6, using high-sensitivity testing for both sample types, saliva is more frequently positive than nasal swabs, shown by bolded black lines (Fig. 3B). Comparison of paired samples between day 6 and day 12 both sample types show highly concordant detection. In a later time interval, between days 12 and 16, nasal swabs are more frequently positive than saliva, shown by bolded green lines (Fig. 3B). The median of peak viral loads is higher in nasal swabs than saliva (Fig. 3C), which is also consistent with literature.^26,28,36^

Many testing strategies and decisions are based on the presence or absence of symptoms.^2,74^ We considered the positivity rate of high- or low-sensitivity testing methods with each sample type during the first ten days of test-positive infection, separating into categories of no symptoms or symptomatic if the participant reported at least one COVID-19-like symptom (Fig. 3D). This interval of 10 days was selected, to capture the pre-symptomatic and symptomatic phases of infection for this cohort, while avoiding samples collected during the post-symptomatic phase of infection. Regardless of symptom status or sample type, high-sensitivity testing results in considerably higher positivity frequency than low-sensitivity testing (Fig. 3D). For samples collected while participants were asymptomatic, high-sensitivity saliva testing was more effective (74% positivity) than nasal swabs of high- (52%) or low-sensitivity (34%) or of saliva of low-sensitivity (17%). In contrast, during symptomatic phases, which is often concurrent with peak nasal viral loads (Fig. 2), high-sensitivity saliva (88%) and high-sensitivity nasal swab testing (89%) have similar positivity rates. Additionally, based on our measured viral loads, low-sensitivity nasal-swab testing is predicted to perform better in symptomatic cohorts than in asymptomatic persons, consistent with how these tests were originally authorized.

## Limitations

Our study needs to be interpreted in the context of its limitations. First, our results capture viral load dynamics from a limited number of individuals from one region of one country with limited SARS-CoV-2 diversity. A larger study with individuals of diverse ages, genetic backgrounds, medical conditions, COVID-19 severity, and SARS-CoV-2 lineages would be ideal to provide a more nuanced and representative understanding of viral dynamics in saliva and nasal-swab samples. Second, the commercial inactivating buffer used here (Spectrum SDNA-1000) is not authorized (at the time of this writing) for the sample collection of nasal swabs. Third, we have paired data for saliva and anterior nares nasal swabs but do not compare nasopharyngeal (NP) swabs, sputum, or other lower-respiratory specimens. We do not know whether other sampling sites, such as nasopharyngeal swabs or oropharyngeal swabs, would have provided earlier or later detection than saliva. As mentioned above, parallel sampling of multiple respiratory sites should be done as new variants emerge. Fourth, we do not have data for low-sensitivity tests or any antigen tests, and are inferring ability to pick up infections based on the quantified viral load in the participant samples and the LODs reported by the FDA for the diagnostic platforms. Fifth, our investigation of SARS-CoV-2 RNA stability in each sample type (saliva, nasal swabs) in the inactivating buffer at 4 °C during storage suggests some degradation may have occurred in some samples, and revealed subtle differences between degradation in saliva and nasal swabs (Fig. S3). See Supplement for a complete analysis of RNA stability. Sixth, our samples were self-collected by participants after detailed training by our study coordinators during the study enrollment process. Samples self-collected without such training may result in lower quality specimens. Similarly, our participants were able to collect saliva samples during specific parts of their day (after waking and before going to bed) without eating, drinking, or brushing teeth prior to collection. This protocol may not be practical in all settings and we do not know how deviations from this protocol would affect viral loads in saliva. Lastly, our samples were self-collected in a guanidinium-based inactivating and stabilizing buffer that preserves viral RNA but eliminated the opportunity to also perform viral culture.

## Conclusions

By rapidly enrolling household members at high risk for contracting COVID-19 and having them self-sample and report symptoms twice daily in paired respiratory sites, we were able to observe patterns in SARS-CoV-2 viral load in the earliest days of infection. All seven participants tested negative by both sample types (saliva and nasal swabs) upon enrollment, ensuring we captured the earliest detectable SARS-CoV-2 viral load (within 12 hours) in both sample types. Our dataset helps inform diagnostic testing strategies by showing differences in viral loads in paired nasal swabs and saliva samples at high temporal resolution (twice daily) during the early days and pre-symptomatic phases of infection.

### We made five conclusions from our study

First, choosing the correct respiratory sampling site is critical for earliest detection of SARS-CoV-2 infection. In our study, alignment of longitudinal data to the first day of positivity clearly shows the superiority of high-sensitivity saliva testing for detection in the first 5.5 days of infection (Fig. 3A, Fig. S5). Although sampling with NP swabs may be considered by some to be the gold-standard for COVID-19 testing, there are no data suggesting NP swabs are superior for earliest detection of SARS-CoV-2 infection. Furthermore, anterior-nares swabs and saliva tests have become more common than NP swabs for practical reasons. Given our data, early infection viral load dynamics in multiple sampling sites should be investigated and compared with saliva as new SARS-CoV-2 variants emerge.

Second, our data explain the conflicting results in the literature comparing the performance of testing from paired respiratory sites, with some studies showing nasal-swabs outperform saliva^26,28,36^ and others showing saliva (or oral fluid) has equivalent or better detection to nasal-swabs.^21,30,37–45^ Through longitudinal rather than cross-sectional sampling, we show the relative viral loads in each respiratory site is a factor of infection stage (shown in time intervals in Fig. 3B), and the kinetics of viral load over the course of the infection may be quite distinct in each sample type for an individual (Fig. 2). Most studies examining paired sample types have enrolled participants after a positive test or after symptom onset; as our data show, detectable viral loads precede symptoms, in most cases (5/7 participants) by several days (Fig. 2). Thus, enrollment after positivity or after symptom onset is not an appropriate study design to determine the respiratory sample type in which the virus is first detectable (Fig. 3A, Fig. S5).

Third, peak viral load measured in nasal swabs (Fig. 3C) is not representative of detectable viral load in the earliest days of infection (Fig. 3A) nor during the pre-symptomatic phase (Fig. 3D). Early in an infection, it is inappropriate to assume that a person is “not infectious” or “has low viral load” based on a measurement from a single sample type such as a nasal swab, given that saliva is known to carry infectious virus.^73^ In our study, we observed a participant with very high (>10^7^-10^8^ copies/mL) viral load in saliva samples while the paired nasal swab was either negative (Fig. 2G, ROI#2) or had low (∼10^3^ copies/mL) viral load (Fig. 2G, day after ROI#2). Quantitative SARS-CoV-2 culture from paired saliva and swab samples is still needed to understand infectiousness during the early stages of SARS-CoV-2 infection.

Fourth, using a diagnostic test with high analytical sensitivity (Fig. 3D), rather than a test of a particular detection method (RT-PCR, antigen, next-generation sequencing, etc.), is essential to early detection. Often the test type (e.g., RT-qPCR) is incorrectly equated with high analytical sensitivity, and some current travel and work guidelines specify a test type (e.g., RT-qPCR) rather than a particular test LOD. However, this is an invalid assumption; FDA testing^17^ demonstrated that sensitivity of RT-qPCR tests ranges from highly sensitive (e.g., LOD of 180 NDU/mL for PerkinElmer and 450 NDU/mL for Zymo Research) to substantially less sensitive (e.g., LOD of 180,000 NDU/mL for TaqPath COVID-19 Combo Kit and 540,000 NDU/mL for Lyra Direct SARS-CoV-2 Assay). FDA’s NDU/mL is approximately equivalent to the copy/mL scale used in this paper. The low-sensitivity end of this RT-qPCR sensitivity range (corresponding to the higher LOD values) overlaps with the range of low-sensitivity rapid isothermal nucleic-acid tests (e.g., LOD of 180,000 NDU/mL for Atila BioSystems and 300,000 NDU/mL for Abbott ID NOW tests) and approaches the analytical sensitivity range of antigen tests. Therefore, to achieve early detection, tests with high sensitivity rather than tests of a particular type should be chosen. With many strategies for asymptomatic screening/surveillance testing in use, it is critically important to consider whether the LODs of the tests would be able to detect early infection, and to prompt actions that minimizes transmission.

Fifth, our data show the utility of combining knowledge of the appropriate respiratory site and the appropriate test analytical sensitivity for achieving earliest detection. Among our unvaccinated participants, when a high-sensitivity test was combined with saliva as a sample type, SARS-CoV-2 infection was detected up to 4.5 days before viral loads in nasal swabs reached the LODs of low-sensitivity tests (Fig. 2F). Although high-sensitivity saliva testing was usually able to detect earlier than nasal swabs (Fig. 3A, Fig. S5), during the peak of the infection viral loads in nasal swabs were usually higher than in saliva (Fig. 3C). Furthermore, SARS-CoV-2 was detected in saliva with high-sensitivity methods and the viral loads were low (Fig. 2, Fig. 3D, Fig. S5); low-sensitivity saliva tests would likely not have been able to detect these infections early. These observations support the preferred use of nasal swabs in environments where only low-sensitivity testing is available, although the performance of such testing for early detection is poor (Fig. 3D). These observations also show that the choice of the optimal respiratory sampling site is nuanced and depends on the phase of the infection being detected (early vs peak) and on the analytical sensitivity of the test being used with each sample type.

Our work suggests three steps to improve effectiveness of diagnostic tests in early detection and preventing transmission of SARS-CoV-2 as new variants emerge and as infections spread to additional segments of the global population: (1) Additional longitudinal studies are needed that include high-frequency collection from multiple respiratory sites using quantitative assays with high analytical sensitivity. (2) Policy makers need to use such quantitative data to revise and optimize testing, surveillance, and screening guidelines to ensure early detection of SARS-CoV-2 infections and reduction of transmission. (3) Innovation is needed to produce rapid point-of-care tests with high analytical sensitivity for a range of sample types (including saliva) at a price point to enable global distribution.

As new SARS-CoV-2 variants emerge, quantitative studies of the kinetics of early-stage viral loads must be continually updated. Importantly, such studies should be undertaken in people of a wide range of ethnicities, races, health conditions, and ages. For example, children under 12 years of age remain ineligible for vaccination even as schools around the country reopen for in-person studies. Given the potential differences in viral kinetics between children and adults, it is critical to collect quantitative viral-load data from children to understand the most effective testing strategies for this population. Quantitative studies of viral-load dynamics must also include vaccinated persons. Breakthrough cases are often asymptomatic^75^ and recent evidence suggests that vaccinated individuals may transmit infections from the new variants, including Delta.^7^ Another reason for continued monitoring of early viral kinetics is that viral evolution, including of host tropism, can markedly diminish the effectiveness of a diagnostic strategy. In one study, decreased clinical sensitivity of NP swabs was observed in SARS-CoV-2 variant B.1.616,^46^ which may indicate a tropism shift of the virus into lower-respiratory compartments. Finally, quantitative data must be acquired in parallel with viral-culture data to understand the viral loads and phases of infection that are most relevant to transmission.

Early detection of infection clearly reduces community transmission, however for most of the COVID-19 pandemic, policy makers have had to develop testing strategies in the absence of quantitative data on viral kinetics from the earliest stage of infection. Testing strategies have been guided by the available viral-load data, which has come mostly from hospitalized patients, symptomatic people, and people who have tested positive with the commonly available tests; but these data cannot inform on the best strategies for *earliest* detection. Moreover, lacking such data-based guidance, diagnostic tests have been used incorrectly (with false-negative results due to using tests with insufficient sensitivity) in several scenarios, resulting in outbreaks that could have been prevented with an appropriate testing strategy.^76–82^ Once armed with data on early viral-load kinetics of new and emerging variants, policy makers will be able to develop targeted testing strategies (frequency of sampling, required test sensitivity, and appropriate respiratory site) for key populations undergoing regular repeated testing, such as hospital staff and nursing-home staff. Such data would also inform optimal testing guidelines for early detection in situations where a single test is typically administered to admit a person into an environment (e.g., persons embarking on a flight or cruise, summer camps, or admission to a country) where the impact of false negatives is particularly high.

One barrier to implementing such more advanced testing strategies is availability of appropriate tests. Because the optimal sample type for early detection might be different for different populations, or might change as new variants emerge, tests with robust high analytical sensitivity across multiple sample types are needed. Developing such tests is challenging because it requires incorporating robust sample-preparation technology to purify and concentrate pathogen nucleic acids from diverse human matrices, from upper respiratory (e.g. fluids from the nasal, nasopharyngeal, oral and oropharyngeal compartments, captured in swabs or saliva) to lower respiratory samples (e.g. sputum, tracheal aspirate, bronchoalveolar lavage). It is even more challenging to incorporate such sample-preparation technology into tests that can be broadly deployed—at very low cost—at the point of care in limited-resource settings (such as schools, homes, and businesses, and especially in the developing countries). Development of such highly sensitive, rapid, and inexpensive tests with broad sample-type compatibility is urgently needed.

We hope our data, important work by others in this area,^20,21,58,73^ and future quantitative studies of early viral-load kinetics will lead to improved testing strategies to combat the current COVID-19 pandemic. The methodology for performing such studies efficiently and quickly will likely be extendable to defining strategies for early detection of causative pathogens in subsequent pandemics.

## Methods

Refer to the Supplementary Information for detailed methods.

### Participant Population

This study is an extension of our previous study examining viral load in saliva.^19^ Both studies were reviewed and approved by the Institutional Review Board of the California Institute of Technology, protocol #20-1026. All participants provided either written informed consent or (for minors ages 6-17) assent accompanied by parental permission, prior to enrollment. Participants were eligible if they had recently (within 7 days) been diagnosed with COVID-19 by a CLIA laboratory test, or lived with someone meeting who had. Demographic and medical information for the seven participants described here can be found in Table S1.

### Questionnaires and Symptom Monitoring

Acquisition of participant data was performed as described in our previous study.^19^ Symptoms (including those listed by the CDC^83^) were reported by participants twice daily in parallel with sample collection. Participants recorded any COVID-19-like symptoms (as defined by the CDC^83^) on a symptom-tracking card or on a custom app run on REDCap.

Participants self-collected nasal-swab (1 swab) and saliva (∼1.5mL) samples in the Spectrum SDNA-1000 Saliva Collection Kit (Spectrum Solutions LLC, Draper, UT, USA), which contains 1.5mL of liquid buffer, at home twice per day (after waking up and before going to bed), per manufacturer’s guidelines. Of note, at the time of this writing, Spectrum devices are not approved for the collection of nasal-swab samples. Participants were instructed not to eat, drink, smoke, brush their teeth, use mouthwash, or chew gum for at least 30 min prior to donating. Prior to nasal-swab donation, participants were asked to gently blow their noses to remove debris. Participants were provided with one of the following types of sterile flocked swabs: Nest Oropharyngeal Specimen Collection Swabs (Cat. NST-202003, Stellar Scientific, Baltimore, MD, USA) Puritan HydraFlock Swab (Cat. 25-3000-H E30, Puritan, Guilford, ME, USA) or Copan USA FLOQSwab (Cat. 520CS01, VWR International, Radnor, PA, USA). Participants were instructed to swab each nostril for four complete rotations using the same swab while applying gentle pressure, then to break the tip of the swab into the Spectrum tube and securely screw on the cap. A parent or legal guardian assisted all minors with swab collection and they were instructed to wear a face covering during supervision.

Samples were stored at 4 °C and were equilibrated to room temperature before being processed with extraction protocols.

### RNA Extraction and Nucleic Acid Quantification

Participant saliva and anterior-nares swab samples were extracted using the KingFisher Flex 96 instrument (ThermoFisher Scientific) with the MagMax Viral Pathogen I Nucleic Acid Isolation kit (Cat. A42352, Applied Biosystems, Waltham, MA, USA) guided by ThermoFisher technical notes for SARS-CoV-2 modification and saliva.

RT-qPCR was performed as previously described^19^ using the CDC 2019-Novel Coronavirus (2019-nCoV) Real-Time RT-PCR Diagnostic Panel,^66^ with duplicate reactions. To establish the limit of detection (LOD) for each sample type (saliva, nasal swab), 20 contrived samples with the equivalent of 1,000 copies/mL were prepared, individually extracted as described above, and subjected to RT-qPCR, with a positive result for detection (as defined in the EUA for the CDC RT-qPCR assay) in ≥ 19 of 20 (≥95%) of replicates establishing the LOD (Fig. S1 A,B). For quantification of viral load from RT-qPCR, a standard curve was prepared for both the saliva and nasal-swab protocols with a serial dilution of known concentration (based on the certificate of analysis, COA) of heat-inactivated SARS-CoV-2 particles (Batch 70034991, Cat. NR-52286, BEI Resources, Manassas, VA, USA) in the inactivating buffer from the Spectrum SDNA-1000 Saliva Collection Kit (Spectrum Solutions LLC, Draper, UT, USA) were prepared in triplicate for each concentration, then extracted and measured by RT-qPCR as described above. For positive samples meeting quality control Cq cutoffs based on the CDC guidelines,^66^ the mean Cq of duplicate positive reactions was used for conversion to viral load using the equations shown below obtained from these calibration curves.

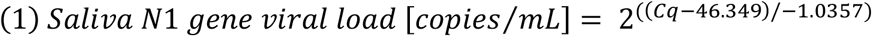

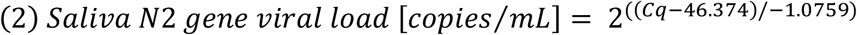

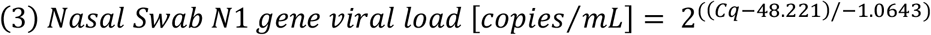

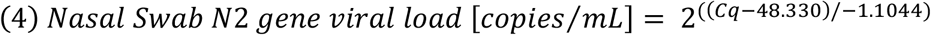

Quantification was also performed by reverse-transcription droplet digital PCR (RT-ddPCR) on elutions from both the calibration curve samples (Fig. 1, Fig. S2) and participant samples (Fig. 1) using the Bio-Rad SARS-CoV-2 Droplet Digital PCR kit (Cat. 12013743, Bio-Rad). Droplets were created using the QX200 Droplet Generator (Cat #1864002, Bio-Rad), thermocycling performed on Bio-Rad C1000 and detected using the QX200 Droplet Digital PCR system (Cat. 1864001, Bio-Rad). Samples were analyzed with QuantaSoft analysis Pro 1.0.595 software following Bio-Rad’s research-use only (RUO) SARS-CoV-2 guidelines.^67^

### Viral Sequencing

Saliva and nasal-swab samples with an *N1* gene Cq of below 26 were sent to Chan Zuckerberg Biohub for SARS-CoV-2 viral genome sequencing, a modification of Deng *et al*. (2020)^84^ as described in Gorzynski *et al*. (2020).^85^ Sequences were assigned pangolin lineages described by Rambaut *et al*. (2020)^86^ using Phylogenetic Assignment of Named Global outbreak LINeages software v2.3.2 (github.com/cov-lineages/pangolin). Chan Zuckerberg Biohub submitted consequence genomes to GISAID.

## Data Availability

Data are available on CaltechDATA at https://data.caltech.edu/records/1942

## DATA AVAILABILITY

Data are available on CaltechDATA at https://data.caltech.edu/records/1942

## ACKNOWLEDGEMENTS

We thank Lauriane Quenee, Junie Hildebrandt, Grace Fisher-Adams, RuthAnne Bevier, Chantal D’Apuzzo, Ralph Adolphs, Victor Rivera, Steve Chapman, Gary Waters, Leonard Edwards, Gaylene Ursua, Cynthia Ramos, and Shannon Yamashita for their assistance and advice on study implementation and/or administration. We thank Jessica Leong, Jessica Slagle, Mika Walton, Angel Navarro, Daniel Brenner, and Ojas Pradhan for volunteering their time to help with this study, Si Hyung Jin for helping with a literature review, and Mary Arrastia for providing biosafety support. We thank Maira Phelps, Lienna Chan, Lucy Li, Dan Lu and Amy Kistler at the Chan Zuckerberg Biohub for performing SARS-CoV-2 sequencing. We thank Angie Cheng, Susan Magdaleno, Christian Kis, Monica Herrera, and Zaina Lemeir for technical discussions regarding saliva extraction and ddPCR detection. We thank Jennifer Fulcher, Debika Bhattacharya and Matthew Bidwell Goetz for their ideas on potential study populations and early study design. We thank Omai Garner and David Beenhouwer for providing materials for initial nasal-swab validation. We thank Martin Hill, Alma Sanchez, Scott Kim, Debbie Noble, Nina Paddock, Whitney Harrison, Emily Holman, Isaac Turner, Vivek Desai, Luke Wade, Tom Mayell, Stu Miller, and Jennifer Howes for their support with recruitment. We thank Allison Rhines, Karen Heichman, and Dan Wattendorf for valuable discussion and guidance. Finally, we thank all the case investigators and contact tracers at the Pasadena Public Health Department and the City of Long Beach Department of Health & Human Services for their efforts in study recruitment and their work in the pandemic response.

## SUPPLEMENTAL INFORMATION

- Figures S1 – S5
- Table S1
- Author Contributions

## Supplemental Information

**Figure S1.**
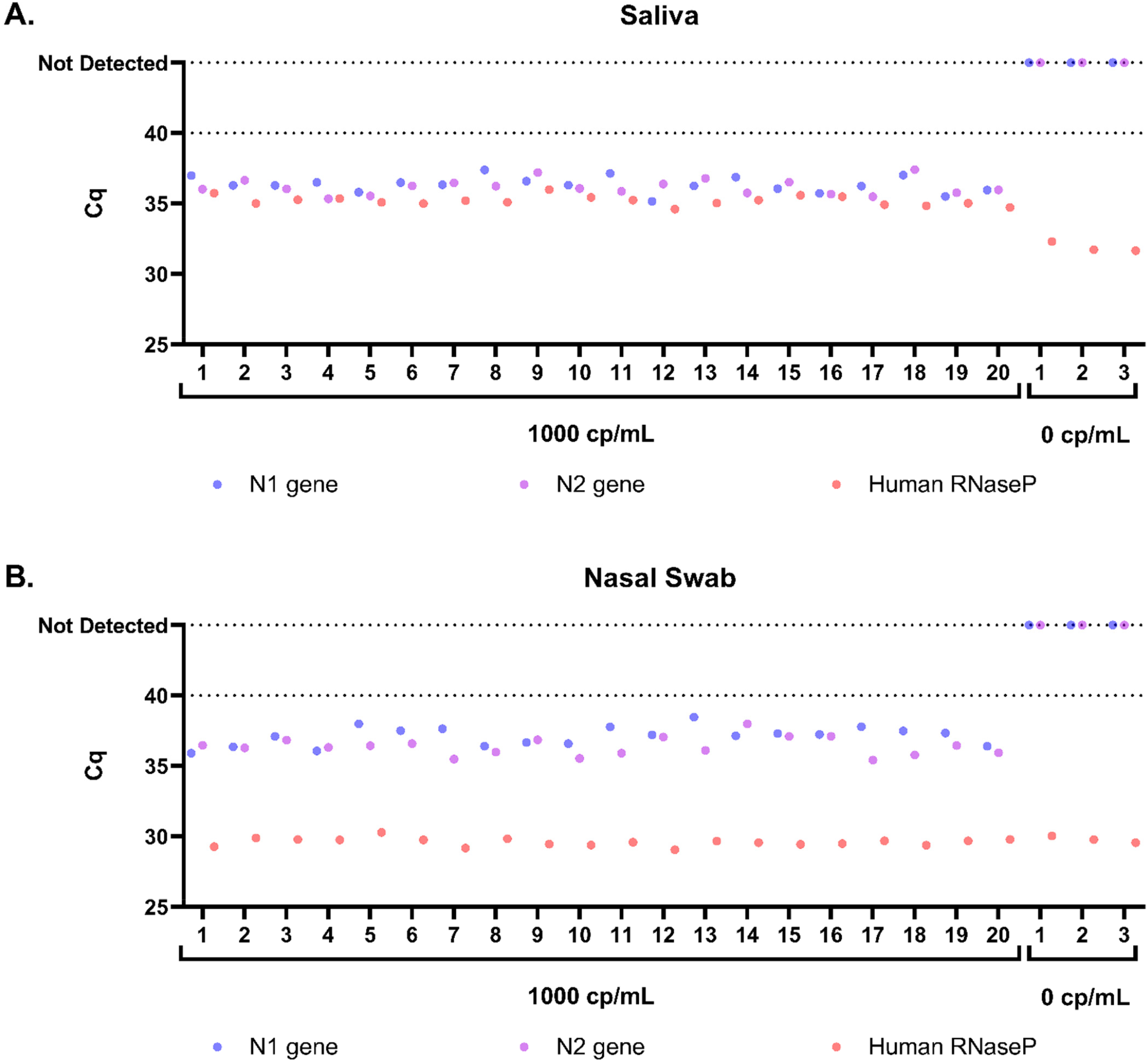
Limit of detection of saliva and nasal-swab RT-qPCR assays used in this study. RT-qPCR quantification cycle (Cq) for SARS-CoV-2 *N1* gene (blue circle), *N2* gene (purple circle), and human *RNase P* gene (orange circle) in 20 replicates of pooled matrix spiked with 1000 copies/mL (cp/mL) heat-inactivated SARS-CoV-2 RNA and 3 replicates of pooled matrix spiked with a buffer blank for saliva (A) and nasal-swab (B) samples. Duplicate RT-qPCR reactions were performed for each extraction replicate and the averages are shown, with the following three exceptions: replicate 9 (saliva), in which the *N1* gene only amplified in 1 of the duplicate runs (*N2* in this run was positive, so per CDC EUA guidelines^87^ this run was interpreted as inconclusive), replicate 10 (nasal swab) in which the *N2* gene only amplified in 1 of the duplicate runs (N1 in this run was positive, so this run was interpreted as inconclusive), and replicate 18 (nasal swab) in which the N1 gene only amplified in 1 of the duplicate runs (*N2* in this run was positive, so this run was also interpreted as inconclusive). None of the samples spiked at 1000 copies/mL gave a negative detection result.

**Figure S2.**
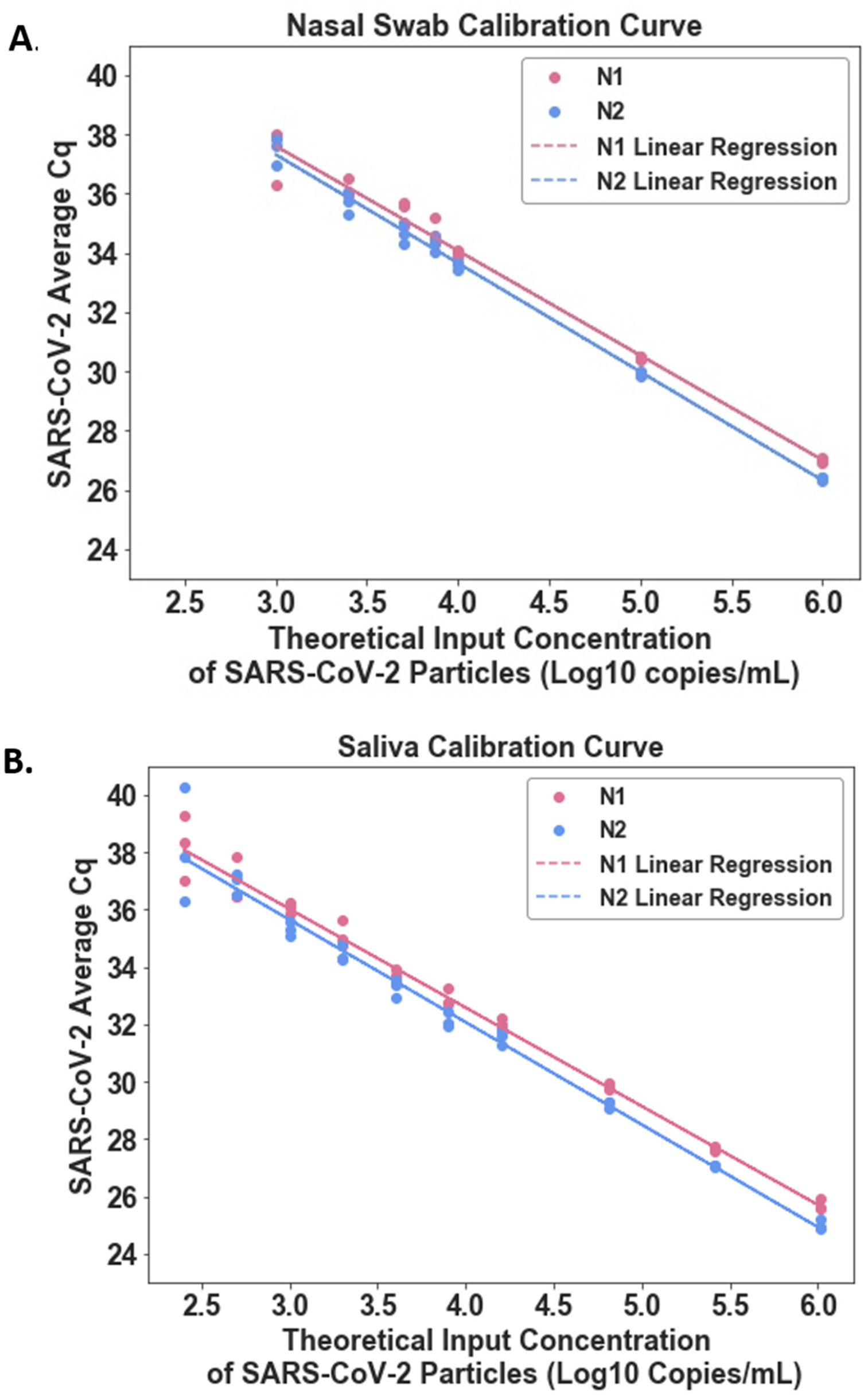
Calibration curve of SARS-CoV-2 inactivated particles to establish viral load conversion equations. Linear regression of RT-qPCR quantification cycle (Cq) for *N1* (red circle) and *N2* (blue circles) genes at known concentrations of inactivated SARS-CoV-2 particles for saliva (A) or nasal swab (B) using this study’s collection and laboratory workflows. Triplicate replicates per concentration were performed. Linear regression for *N1* represented by red line and *N2* represented by blue line. Linear regression R^2^ values are 0.986 for *N1* in nasal swabs, 0.994 for *N2* in nasal swabs, 0.989 for *N1* in saliva, and 0.979 for *N2* in saliva.

**Figure S3.**
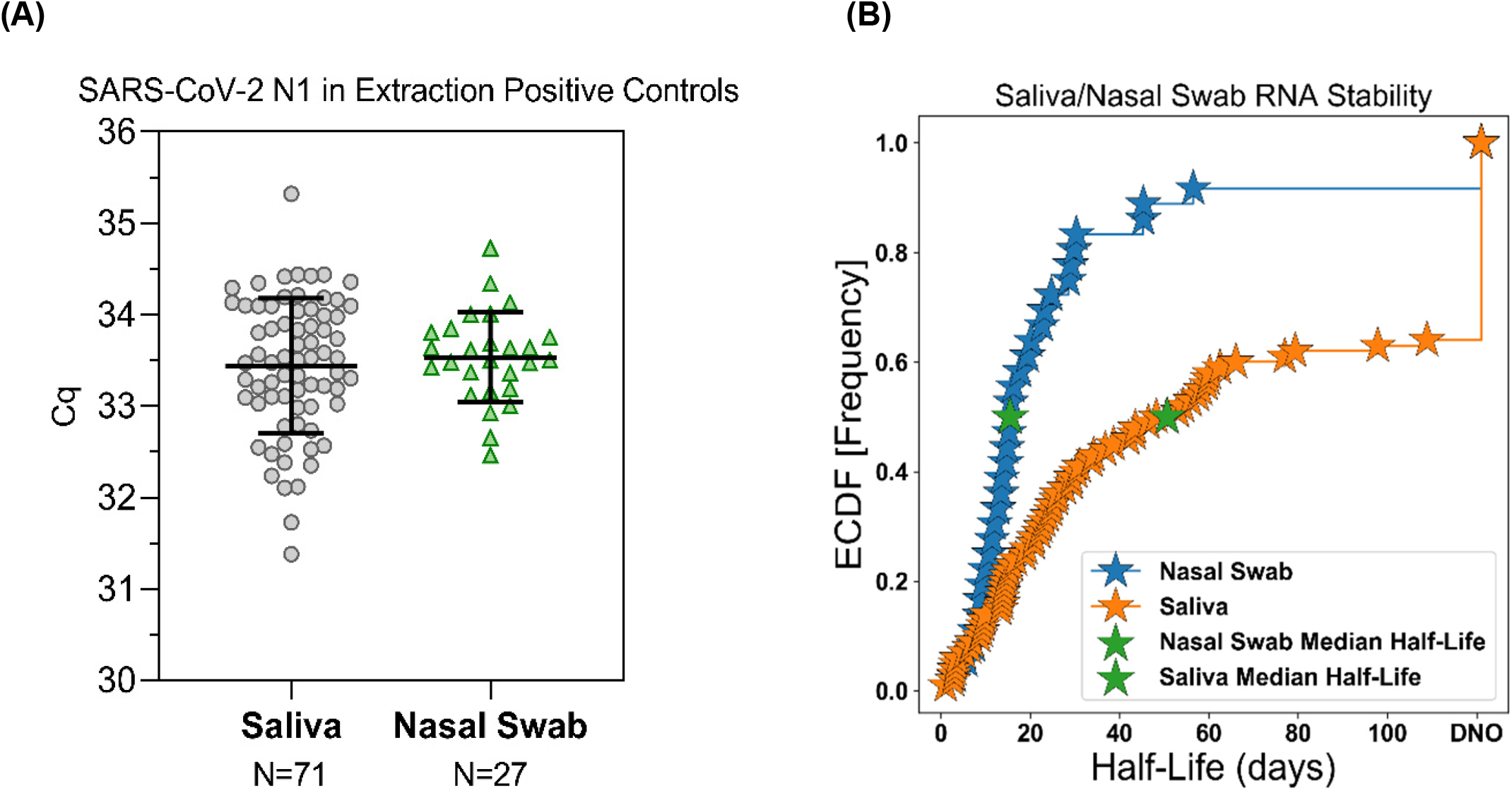
SARS-CoV-2 RNA stability over time in Spectrum SDNA-1000 buffer at 4 °C. (A) Positive extraction control samples from 71 saliva extraction runs and 27 nasal-swab extraction runs are included to show the measurement noise in the quantification workflow. The standard deviation for the positive control measurements was 0.74 Cq for saliva and 0.49 for nasal swab. (B) Empirical cumulative distribution functions (ECDF) for the observed half-life (days) of participant saliva (orange stars) and nasal-swab (blue stars) samples in Spectrum SDNA-1000 buffer stored at 4 °C. Individual samples were extracted at multiple time points. The ECDF represents the summed probability mass function over the Cq half-lives observed. Half-life in this context refers to the time required to observe a 1 Cq increase (representing a 2-fold decrease) in RNA detected by RT-qPCR. The median point is identified with a green star for each sample, at 15.0 days for nasal swabs and 51.0 days for saliva. Of the 110 total participant saliva samples plotted in panel B, 36 samples had no evidence of degradation (DNO) under the time frame measured. Only 3 of the 36 total participant nasal-swab samples plotted in panel B had no evidence of degradation (DNO) under the time frame measured. DNO = degradation not observed, meaning that the difference in extraction Cq values of the same sample at multiple time points was within 1 standard deviation observed in replicate extraction positive controls for the respective sample type, as shown in panel A.

**Figure S4.**
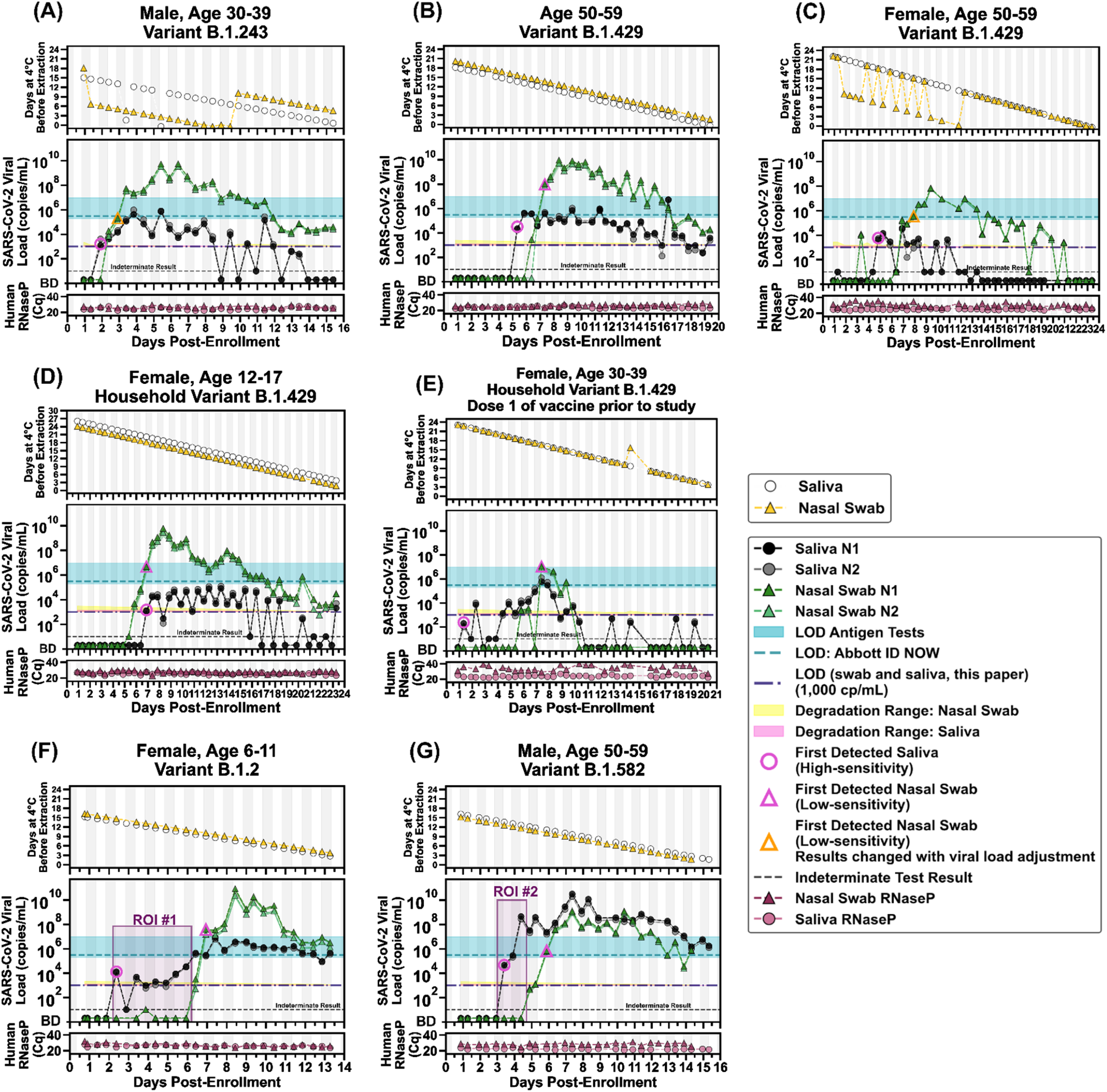
Predicted impact of SARS-CoV-2 RNA stability on viral loads shown in Fig. 2. (A-G) The time [days] of sample storage at 4 °C between sample collection and RNA extraction is shown in the topmost panels. Open circles represent saliva samples and yellow triangles represent the nasal swabs. Viral load (black and grey circles) calculations are corrected for the median half-life (1 Cq decrease in RNA detected by RT-qPCR) of each sample type and the duration of storage at 4 °C before quantification (15 days for 2-fold decrease in detected RNA in nasal swabs and 51 days for 2-fold decreased in detected RNA in saliva). The degradation ranges, represented by a shaded yellow (nasal swab) or pink (saliva) region to represent how a measured value of 1,000 copies/mL may have degraded from concentrations in this range. As in Fig. 2, ND = not detected for Cqs ≥40 (see Methods for details). The limit of detection (LOD) of the saliva and nasal-swab assays used here (1,000 cp/mL) is indicated with the purple dashed line; the LOD of the Abbott ID NOW (300,000 NDU/mL^17^) is indicated by the horizontal green dashed line; the range of LODs of antigen tests (horizontal green bar) are shown for reference (data are from Table S2 in ref. ^19^). A diagnostic test does not provide reliable detection for samples with viral loads below its LOD. For each participant, the first detected saliva point is emphasized with a pink circle and their first nasal-swab point above the LOD of the ID NOW is emphasized with a pink triangle. Vertical shading in grey indicates nighttime (8pm – 8am). Internal control of *RNase P* gene Cqs from the CDC primer set are provided for each sample to compare self-sampling consistency and sample integrity (failed samples, where *RNase P* Cq ≥40, are not plotted). Samples with an indeterminate result by the CDC RT-qPCR assay are shown along the horizontal black dashed line. Participant gender, age range, and SARS-CoV-2 variant is given in each panel’s title. Two regions of interest (ROI) are indicated by purple-shaded rectangles and discussed in the main text. For the two points that change interpretation with the viral load adjustment, orange triangles show which new data points become the first nasal-swab point in range of low-sensitivity tests.

**Figure S5.**
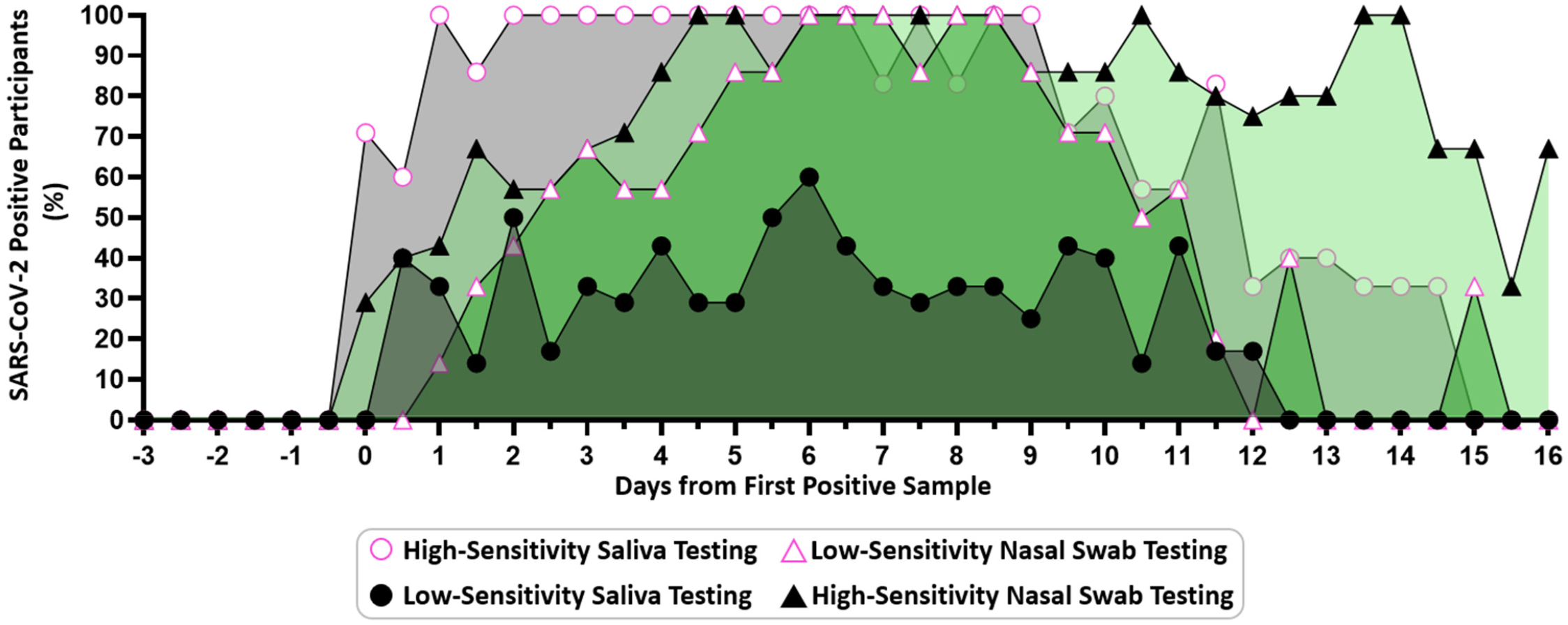
Participants positive by each sample type and test sensitivity at each timepoint from the incidence of infection. Infection time courses were aligned to the first positive result from our high-sensitivity (LOD of ≤1e3 copies/mL) testing for either sample type. Using the individual SARS-CoV-2 *N1* viral load values for each participant individually shown in Figure 2, we show the percentage of participants who were detected by our high-sensitivity saliva (black circle with pink border) or nasal swab (green triangle) assay (LOD ≤1e3 copies/mL) or could be inferred to be detectable by a low-sensitivity nasal swab (green triangle with pink border) or saliva (black circle) test (with an LOD of 1.9e5 copies/mL) at a given timepoint. Positivity by the high-sensitivity saliva test and low-sensitivity nasal swab test is also shown in Fig. 3 panel A.

**Table S1.** Study participant demographic data. Figure 2 shows viral loads and symptoms data for the seven participants for whom we observed transmission during their enrollment in the study.

|  | **Age Range**<br>**(Years)** | **Sex** | **Race;**<br>**Ethnicity** | **Reported Medical Conditions Associated**<br>**with Increased Risk of Severe**<br>**COVID-19<sup>88</sup>** |
| --- | --- | --- | --- | --- |
| Fig. 2A, Fig. S4A | 30-39 | Male | Other;<br>Mexican/Mexican-American/Chicano<br>(Salvadoran) | Diabetes |
| Fig. 2B, Fig. S4B | 50-59 | Male | Do not wish to respond;<br>Mexican/Mexican-American/Chicano | None |
| Fig. 2C, Fig. S4C | 50-59 | Female | White;<br>Mexican/Mexican-American/Chicano<br>(Spanish-American from<br>Spain) | None |
| Fig. 2D, Fig. S4D | 12-17 | Female | White;<br>Mexican/Mexican-American/Chicano | None |
| Fig. 2E, Fig. S4E | 30-39 | Female | White;<br>Mexican/Mexican-American/Chicano | None |
| Fig. 2F, Fig. S4F | 6-11 | Female | White;<br>Non-Hispanic | None |
| Fig. 2G, Fig. S4G | 50-59 | Male | American Indian or<br>Alaskan Native, White;<br>Other Hispanic, Latinx or<br>Spanish origin | Obesity |

### Supplementary Methods

#### Participant Population

This study is an extension of our previous study examining viral load in saliva.^19^ Both studies were reviewed and approved by the Institutional Review Board of the California Institute of Technology, protocol #20-1026. All participants provided either written informed consent or (for minors ages 6-17) assent accompanied by parental permission, prior to enrollment. Household index cases were eligible for participation if they had recently (within 7 days) been diagnosed with COVID-19 by a CLIA laboratory test. Individuals were ineligible if they were hospitalized or if they were not fluent in either Spanish or English. All participant data were collected and managed using REDCap (Research Electronic Data Capture) on a server hosted at the California Institute of Technology. Demographic and medical information for the seven participants described here can be found in Table S1.

#### Questionnaires and Symptom Monitoring

Acquisition of participant data was performed as described in our previous study.^19^ Briefly, upon enrollment each participant completed an online questionnaire regarding demographics, health factors, prior COVID-19 tests, COVID-19-like symptoms since February 2020, household infection-control practices, and perceptions of COVID-19 risk. Participants also filled out a post-study questionnaire in which they documented medications taken and their interactions with each household member during their enrollment.

Information on symptoms was collected twice daily in parallel with sample collection. Participants recorded any COVID-19-like symptoms (as defined by the CDC^83^) they were experiencing at the time of sample donation on a symptom-tracking card or on a custom app run on REDCap. Whenever possible, participants indicated the self-reported severity of each symptom. Participants were also given the opportunity to write-in additional symptoms or symptom details not otherwise listed.

#### Collection of Respiratory Specimens

Participants self-collected both their nasal-swab and saliva samples using the Spectrum SDNA-1000 Saliva Collection Kit (Spectrum Solutions LLC, Draper, UT, USA), which contains 1.5 mL of liquid buffer, at home twice per day (after waking up and before going to bed), per manufacturer guidelines. Of note, at the time of this writing, Spectrum devices are not approved for the collection of nasal-swab samples. Participants self-collected nasal-swab (1 swab) and saliva (∼1.5mL) samples in the Spectrum SDNA-1000 Saliva Collection Kit (Spectrum Solutions LLC, Draper, UT, USA), which contains 1.5mL of liquid buffer, at home twice per day (after waking up and before going to bed), per manufacturer’s guidelines. Of note, at the time of this writing, Spectrum devices are not approved for the collection of nasal-swab samples.

Participants were instructed not to eat, drink, smoke, brush their teeth, use mouthwash, or chew gum for at least 30 min prior to donating. Prior to nasal-swab donation, participants were asked to gently blow their noses to remove debris. Participants were provided with one of the following types of sterile flocked swabs: Nest Oropharyngeal Specimen Collection Swabs (Cat. NST-202003, Stellar Scientific, Baltimore, MD, USA) Puritan HydraFlock Swab (Cat. 25-3000-H E30, Puritan, Guilford, ME, USA) or Copan USA FLOQSwab (Cat. 520CS01, VWR International, Radnor, PA, USA). Participants were instructed to swab each nostril for four complete rotations using the same swab while applying gentle pressure, then to break the tip of the swab into the Spectrum tube and securely screw on the cap. A parent or legal guardian assisted all minors with swab collection and they were instructed to wear a face covering during supervision. Tubes were labeled and packaged by the participants and transported at room temperature by a touch-free medical courier to the California Institute of Technology daily for analysis.

Upon receipt of the samples in the California Institute of Technology laboratory, each sample was inspected for quality. A sample failed quality control if the preservation buffer was not released from the Spectrum SDNA-1000 cap, or if sample tubes were leaking or otherwise unsafe to handle. Samples that failed quality control were not processed. Inactivated samples were stored at 4 °C and were equilibrated to room temperature before being processed with extraction protocols.

#### RNA Extraction Protocols

Participant saliva and anterior-nares swab samples were extracted using the KingFisher Flex 96 instrument (ThermoFisher Scientific) with the MagMax Viral Pathogen I Nucleic Acid Isolation kit (Cat. A42352, Applied Biosystems, Waltham, MA, USA) guided by ThermoFisher technical notes for SARS-CoV-2 modification and saliva. Each extraction batch, depending on the sample type being extracted, contained a contrived SARS-CoV-2 negative control sample containing either 225 µL of Spectrum buffer mixed with 225 µL of commercial pooled human saliva (Lee Bio 991-05-P-PreC) or 240 µL of Spectrum buffer with 10 µL of pooled commercial nasal fluid (Lee Bio 991-13-P-PreC); a contrived SARS-CoV-2 positive control sample was also included in each extraction batch, with the formulations above, but with the Spectrum buffer spiked with 7,500 genomic copy equivalents/mL of heat-inactivated SARS-CoV-2 particles (BEI NR-52286).

Saliva and anterior-nares swab samples were prepared for purification by transferring 550 µl (for saliva) or 250 µl (for nasal swab) of each sample from its corresponding Spectrum buffer tube into a 1.5 mL lo-bind Eppendorf tube containing 10 µl (for saliva) or 5 µl (for nasal swab) of proteinase K. To maximize recovery of RNA off swabs, prior to transfer, pipet mixing was performed 5-7 times near the swab in the Spectrum tube before aliquoting into an Eppendorf tube. Saliva samples were vortexed for 30 sec in the Eppendorf tube. Samples were incubated at 65 °C for 10 min, then centrifuged at 13,000 x g for 1 min. Aliquots of 400 µl (for saliva) or 200 µl (for nasal swab) were transferred into a KingFisher 96 deep well plate (Cat. 95040450, ThermoFisher Scientific) and processed following KingFisher protocols MVP_400ul_3washes.bdz (for saliva) or MVP_200ul_2washes.bdz (for nasal swab). Ethanol washes were performed with 80% ethanol. Both sample types were eluted into 100 µl of MagMax viral pathogen elution buffer.

#### RT-qPCR

Quantification of SARS-CoV-2 was performed as previously described.^19^ Briefly, the CDC^66^ SARS-CoV-2 *N1* and *N2* gene primers and probes with an internal control targeting *RNase P* gene primer and probe were run in a multiplex RT-qPCR reaction using TaqPath 1-Step Rt-qPCR Mastermix (Cat. A15299, ThermoFisher Scientific). Reactions were run in duplicate on a CFX96 Real-Time Instrument (Bio-Rad Laboratories, Hercules, CA, USA).

#### RT-ddPCR

Reverse-transcription droplet digital PCR (RT-ddPCR) was performed using the Bio-Rad SARS-CoV-2 Droplet Digital PCR kit (Cat. 12013743, Bio-Rad). Swab samples were processed following the manufacturer’s RUO protocol with 5.5 µl template per 22 µl reaction. A total of 42 participant nasal-swab samples were characterized by RT-ddPCR. Modifications were made for saliva samples by reducing the template addition to 2.75 µl per 22 µl reaction. A total of 63 participant saliva samples were characterized by RT-ddPCR. Prior to adding template, samples were diluted into digital range using nuclease-free water. Droplets were created using the QX200 Droplet Generator (Cat #1864002, Bio-Rad), thermocycling performed on Bio-Rad C1000 and detected using the QX200 Droplet Digital PCR system (Cat. 1864001, Bio-Rad). Samples were analyzed with QuantaSoft analysis Pro 1.0.595 software following Bio-Rad’s RUO SARS-CoV-2 guidelines.^67^

#### Viral Load Standard Curves

A standard curve was prepared for both the saliva and nasal-swab protocols. Samples were prepared with known concentrations (based on the certificate of analysis, COA) of heat-inactivated SARS-CoV-2 particles (Batch 70034991, Cat. NR-52286, BEI Resources, Manassas, VA, USA) in the inactivating buffer from the Spectrum SDNA-1000 Saliva Collection Kit (Spectrum Solutions LLC, Draper, UT, USA). To prepare the samples for the saliva protocol (Fig. S2A) a dilution curve of heat-inactivated SARS-CoV-2 particles (Batch 70034991, Cat no. NR-52286, BEI) in the Spectrum device inactivation buffer at concentrations of 0 copies/mL, 1,000 copies/mL, 2,000 copies/mL, 4,000 copies/mL, 8,000 copies/mL, 16,000 copies/mL, 64,000 copies/mL, 256,000 copies/mL, 1,020,000 copies/mL, and 4,100,000 copies/mL. Samples were made by mixing 620 µL of each concentration of the dilution series with 620 µL of healthy pooled human saliva (Cat, 991-05-P, Lee Biosolutions, Maryland Heights, MO, USA). Triplicate extractions were performed according to the saliva RNA extraction protocol. Each extraction was run in triplicate qPCR reactions and single RT-ddPCR reactions.

To prepare the samples for the nasal-swab RNA extraction protocol (Fig. S2B) we created dilution curves for each sample type (saliva and nasal swab) and each target (*N1* and *N2* genes) using heat-inactivated SARS-CoV-2 particles (Batch 70034991, Cat no. NR-52286, BEI) in the Spectrum inactivation buffer. We ran a dilution series of commercial quantified stock (3.75×10^8^ GE/mL) in a 10-fold dilution series from 1×10^6^ to 1×10^4^ copies/mL with finer resolution down to our LOD at 1×10^3^ copies/mL. To each dilution we added 32 µL of healthy human nasal fluid (Cat No 991-13-P-PreC, Lee Biosolutions) to 768 µL of each dilution for a total volume of 800 µL. Triplicate extractions were performed according to the nasal-swab RNA extraction protocol (described above). Each extraction was run in triplicate qPCR reactions and single RT-ddPCR reactions.

Equations from the calibration curves are below. These calibration curves are used to convert the Cq values obtained by RT-qPCR to viral load in each participant sample. For saliva, viral load is a calculation of viral copies/mL in the saliva corrected for dilution with the Spectrum buffer. We assumed that participants donate saliva to the fill line, matching the1:1 dilution in Spectrum buffer recreated when preparing contrived samples for the saliva calibration curve. For nasal swabs, viral load is a calculation of the concentration of viral copies/mL released from the swab into the 1.5 mL of inactivating buffer (which is a similar volume as the 1-3 mL of viral transport media typically used for sample collection). Concentrations higher than 1,000,000 copies/mL could not be characterized due to a limitation of the available stock concentration of commercial inactivated SARS-CoV-2. To validate linear conversion was acceptable at concentrations higher than 1,000,000 copies/mL, we compared RT-ddPCR and RT-qPCR quantification on some participant samples (Fig. 1).

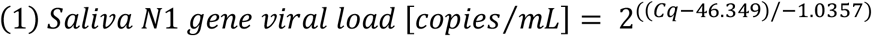

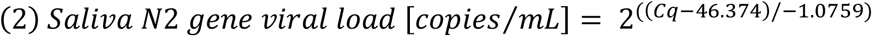

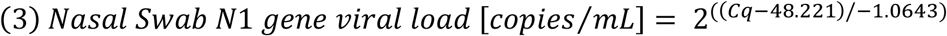

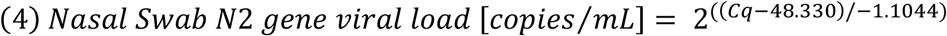

#### Establishment of Limit of Detection

Results of the calibration curve (Fig. S2 A,B) demonstrated 3 of 3 replicates detected at 1,000 copies/mL saliva (for saliva) and 1,000 copies/mL buffer (for nasal swabs). For each sample type (saliva, nasal swab), 20 contrived samples with the equivalent of 1,000 copies/mL were prepared as described above, individually extracted as described above, and subjected to RT-qPCR as described above. The LOD for each sample type through the workflow was considered established if a positive result for detection (as defined in the EUA for the CDC RT-qPCR assay) was obtained for ≥ 19 of 20 (≥95%) of replicates at the input concentration (Fig. S1 A,B).

Three of three replicate sample extractions included in the calibration curves for both contrived nasal-swab samples and contrived saliva samples spiked with heat-inactivated SARS-CoV-2 particles at a concentration of 1,000 copies/mL were detected by RT-qPCR, prompting testing of additional 20 replicates of each sample type spiked at that concentration, individually extracted, and tested by RT-qPCR to establishment of the LOD for our RT-qPCR assay. For both sample types (saliva and nasal swabs), 20 of 20 replicates were positive for SARS-CoV-2 (Fig. S1 A,B), establishing 1,000 copies/mL of saliva and 1,000 copies/mL of swab buffer as the high-sensitivity LOD for our RT-qPCR assays.

#### Data Analysis

Before we converted Cq values to viral load, we used Cq cutoffs based on the CDC guidelines^66^ to exclude from the viral-load plots any points that were indeterminate or fails, and any samples whose RNase P Cq values ≥40. Because we ran duplicate RT-qPCR reactions, the mean Cq of positive reactions was used for conversion to viral load.

#### RNAseq

Saliva and nasal-swab samples below *N1* Cq of 26 were sent to Chan Zuckerberg Biohub for SARS-CoV-2 viral genome sequencing, a modification of Deng et al. (2020)^84^ as described in Gorzynski et al. (2020).^85^ Sequences were assigned pangolin lineages described by Rambaut *et al*. (2020)^86^ using Phylogenetic Assignment of Named Global outbreak LINeages software v2.3.2 (github.com/cov-lineages/pangolin). Consequences viral genomes were submitted to GISAID by Chan Zuckerberg Biohub, see data availability section for accession id details.

#### SARS-CoV-2 RNA Stability at 4 °C

As described above, each extraction batch included a contrived sample spiked with SARS-CoV-2 heat-inactivated particles. For all available saliva or nasal-swab extraction batches, the Cq value of the SARS-CoV-2 *N1* gene in the contrived SARS-CoV-2 positive extraction control was collected. The standard deviation of these measurements was calculated and used to establish a threshold for expected noise between repeat extractions of the same sample. To assess samples for evidence of SARS-CoV-2 RNA degradation, any participant sample that had more than one extraction replicate performed were analyzed. Samples where the difference in Cq values between the extractions was less than the threshold of expected noise between replicate extractions were defined as degradation not observed, (DNO). For samples where the difference was above this threshold, the time for 1 Cq increase (2-fold decrease) in RNA detected by RT-qPCR is described by the term half-life, which was calculated according to Equation 5, below:

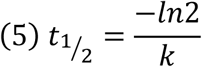

Where “k” is defined as the slope of the linear regression of the natural logarithm of the viral load vs. extraction date (relative to sample collection date). Empirical cumulative distribution functions (ECDF) were calculated using the statsmodels Python package. Confidence intervals were constructed using 2500 bootstrap trials. The median over the entire dataset (saliva or swab) was used as a point estimate of RNA half-life.

Calculations that predict the impact of storage time at 4 °C and RNA stability on viral load are calculated according to Equation 6, below.

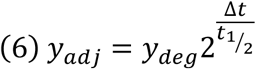

Where y_adj_ is defined as the adjusted viral load, y_deg_ is defined as the viral load before adjustment for degradation (as calculated by Equations 1–4), and t_1/2_ is defined as the RNA half-life, shown in Equation 5.

## Author Contributions

### Listed alphabetically by last name

Reid Akana (RA): Assisted in literature analysis with ES, MKP, AW, MC; collaborated with AW in creating digital participant symptom surveys; assisted with data quality control/curation with JJ, NWS, NS; collaborated with ES, JJ to write data analysis/visualization code; created current laboratory information management system (LIMS) for specimen logging and tracking. Creation of iOS application for sample logging/tracking. Configured a SQL database for data storage. Created an Apache server and websites to view study data. Configured FTPS server to catalog PCR data. Wrote a Python package to access study data. Worked with ES, AW, AR to implement logic that prioritized specimen extraction order. Collaborated with ES, MKP, AW, AR in analyzing RNA stability. Created supplementary figures 3B and 3C.

Jacob T. Barlow (JTB): Created initial specimen tracking database to aid in specimen logging and tracking. Maintenance of database and implementation of corrections. Feedback on manuscript draft.

Alyssa M. Carter (AMC): Received and logged specimens, and performed sample QC. Prepared reagents for and assisted with RNA extractions. Performed RT-qPCR and analyzed RT-qPCR data for both time series and screening experiments. Performed some of the initial experiments that assessed RNA stability in nasal swab samples

Matthew M. Cooper (MMC): Collaborated with AW, MF, NS, YG, RFI, on study design and recruitment strategies. Co-wrote initial IRB protocol and informed consent with AW and NS; assisted in the writing of the enrollment questionnaire; developed laboratory sample processing workflow for saliva with AW and AER; performed sample processing on subset of samples; funding acquisition; collaborated with AR to write data processing/visualization code for observing household transmission events for active study participants. Contributor to the design of the calibration curve for saliva LOD experiments.

Matthew Feaster (MF): Co-investigator; collaborated with AW, MMC, NS, YG, RFI on study design and recruitment strategies; provided guidance and expertise on SARS-CoV-2 epidemiology and local trends.

Ying-Ying Goh (YYG): Co-investigator; collaborated with AW, MMC, NS, MF, RFI on study design and recruitment strategies; provided guidance and expertise on SARS-CoV-2 epidemiology and local trends.

Rustem F. Ismagilov (RFI): Co-investigator; collaborated with AW, MMC, NS, MF, YYG on study design and recruitment strategies; provided leadership, technical guidance, oversight, and was responsible for obtaining funding for the study.

Jenny Ji (JJ): Contributed to study design and study organization and implementation with NS and JAR; co-wrote enrollment questionnaire with NS and AW. Major contributor to curation of participant symptom data. Provided quality control of participant data with RA, NS, NWS. Major contributor to the symptom data analysis and visualization shown in Fig. 2.

Michael K. Porter (MKP): Performed specimen logging and QC, RNA extractions, RT-qPCR, data processing. Performed data acquisition and analysis for and made Figure S1 with AW. Prepared participant sample collection materials and helped with supplies acquisition. Assisted in literature analysis with ES, RA, AW.

Jessica A. Reyes (JAR): Study coordinator; collaborated with NS, AW, NWS, and RFI on recruitment strategies, translated study materials into Spanish, co-wrote informational sheets with AW and NS; created instructional videos for participants; enrolled and maintained study participants with NS and NWS.

Anna E. Romano (AER): Developed laboratory swab sample processing workflow with ES. Optimized extraction and ddPCR protocols working with vendor scientists. Created budgets and managed, planned, and purchased reagents and supplies; developed and validated method for RT-qPCR and RT-digital droplet PCR analysis for saliva & swab samples with MMC, and AW. Performed specimen logging and QC, RNA extractions, RT-qPCR and RT-digital droplet PCR; Design of saliva calibration curve experiment. Analyzed ddPCR data for participant and calibration curve data included in Figure 1. Interpretation of sequence data with AW. Prepared Figure 1 and SI Figure S2 with ES and AW; Collaborated with ES and RA to generate and curate data for RNA stability analysis. Managing logistics for the expansion of the BSL-2+ lab space with ESS. Edited manuscript.

Emily S. Savela (ESS): Coordinated the laboratory team and division of lab work, coordinated lab schedules to ensure completion of time-sensitive analyses of participant samples while complying with COVID-19 lab occupancy restrictions and biosafety requirements. Performed initial nasal-swab workflow validation experiments with AER. Major contributor to workflow validation, methods, biosafety SOPs, and sample storage. Developed a plan for, and executed, the long term sample storage for efficient, safe, storage. Performed specimen logging and QC, RNA extractions, RT-qPCR, data processing, and conducted biosafety training. Performed the data curation and data analysis for Figure 2. Made Figure 2. Minor contributor to symptoms data analysis and visualization with JJ and RA for Figure 2. Experimental design and RNA extractions of the samples, to Figure 1A and minor contributor to Figure S2A with AER. Managing logistics for the expansion of the BSL-2+ lab space with AER and biohazardous waste pickups. Collaborated with ES, RA, and AW to generate data for and curated data set to assess viral RNA stability (Fig. S3). Prepared Fig. S4. Co-wrote the manuscript. Verified the underlying data with AW.

Noah W. Schlenker (NWS): Study coordinator; collaborated with NS, AW, JAR, and RFI on recruitment strategies; enrolled and maintained study participants with NS and JAR; study-data quality control, curation and archiving with RA, JJ, and NS.

Natasha Shelby (NS): Study administrator; collaborated with AW, MMC, RFI, YG, MF on initial study design and recruitment strategies; co-wrote IRB protocol and informed consent with AW and MMC; co-wrote enrollment questionnaire with AW and JJ; co-wrote participant informational sheets with AW and JAR; enrolled and maintained study participants with JAR and NWS; study-data quality control, curation and archiving with RA, JJ, and NWS; reagents and supplies acquisition; assembled Table S1; managed citations and reference library; co-wrote and edited the manuscript.

Colten Tognazzini (CT): Coordinated the recruitment efforts at PPHD with case investigators and contact tracers; provided guidance and expertise on SARS-CoV-2 epidemiology and local trends.

Alexander Winnett (AW): Collaborated with MMC, NS, RFI, YG, MF on initial study design and recruitment strategies; co-wrote IRB protocol and informed consent with MMC and NS; co-wrote enrollment questionnaire with NS and JJ; co-wrote participant informational sheets with NS and JAR and digital survey; developed and validated methods for saliva and nasal-swab sample collection; developed and validated methods for RT-qPCR and RT-digital droplet PCR analysis for saliva and swab samples with AER, ESS, MMC; reagents and supplies acquisition; funding acquisition; developed laboratory sample processing workflow with AER, ESS, and MMC; performed specimen logging and QC, nucleic acid extraction, RT-qPCR, data processing – including experimental data generation for saliva calibration curve (Fig. 1, Fig. S2) designed with MMC and AER, establishment of nasal swab limit of detection (Fig. S1), and viral load timeseries data (Fig. 2) with ESS, AER, MKP, and AMC; interpreted sequencing data with AER; analyzed viral load timeseries data to visualize trends (Fig. 3, Fig. S4) with ESS; generated, analyzed and visualized data to assess degradation of viral RNA in saliva and nasal swab samples with RA, ESS, and AER (Fig. S3); literature analysis with RA, ESS, and MKP; co-wrote sections of the manuscript outlined by ESS and RFI, edited the manuscript. Verified the underlying data with ESS.

